# Association of maternal prenatal copper concentration with gestational duration and preterm birth: a multi-country meta-analysis

**DOI:** 10.1101/2022.08.27.22278198

**Authors:** Nagendra K. Monangi, Huan Xu, Yuemei Fan, Rasheda Khanam, Waqasuddin Khan, Saikat Deb, Jesmin Pervin, Joan T. Price, Lovejeet Kaur, INTERBIO-21^st^ Study Consortium, Abdullah Al Mahmud, Le Quang Thanh, Angharad Care, Julio A. Landero, Gerald F. Combs, Elizabeth Belling, Joanne Chappell, Jing Chen, Fansheng Kong, Craig Lacher, Salahuddin Ahmed, Nabidul Haque Chowdhury, Sayedur Rahman, Furqan Kabir, Imran Nisar, Aneeta Hotwani, Usma Mehmood, Ambreen Nizar, Javairia Khalid, Usha Dhingra, Arup Dutta, Said Mohamed Ali, Fahad Aftab, Mohammed Hamad Juma, Monjur Rahman, Tahmeed Ahmed, M Munirul Islam, Bellington Vwalika, Patrick Musonda, Ulla Ashorn, Kenneth Maleta, Mikko Hallman, Laura Goodfellow, Juhi K. Gupta, Ana Alfirevic, Susan K. Murphy, Larry Rand, Kelli K. Ryckman, Jeffrey C. Murray, Rajiv Bahl, James A. Litch, Courtney Baruch-Gravett, Shailaja Sopory, Uma Chandra Mouli Natchu, Pavitra V Kumar, Neha Kumari, Ramachandran Thiruvengadam, Atul Kumar Singh, Pankaj Kumar, Zarko Alfirevic, Abdullah H. Baqui, Shinjini Bhatnagar, Jane E. Hirst, Cathrine Hoyo, Fyezah Jehan, Laura Jelliffe-Pawlowski, Anisur Rahman, Daniel E. Roth, Sunil Sazawal, Jeffrey S. A. Stringer, Per Ashorn, Ge Zhang, Louis J. Muglia

## Abstract

**Background:** Copper (Cu), an essential trace mineral regulating multiple actions of inflammation and oxidative stress, has been implicated in risk for preterm birth (PTB). We aimed to determine the association of maternal plasma/serum Cu concentrations during pregnancy with PTB risk and gestational duration in a large multi-cohort study including diverse populations.

**Methods:** Gestational duration data and maternal plasma or serum samples of 10,449 singleton live births were obtained from 18 geographically diverse study cohorts. Maternal plasma or serum Cu concentrations were determined by inductively coupled plasma mass spectrometry (ICP-MS) analysis. The associations of maternal Cu with PTB and gestational duration were analyzed using logistic and linear regressions for each cohort. The estimates were then combined using meta-analysis. Associations between maternal Cu and acute phase reactants (APRs), malaria, and HIV infection were analyzed in 1239 samples from the Malawi cohort.

**Findings:** The maternal prenatal Cu concentration in our study samples followed a normal distribution with a mean of 1.92 μg/ml and a standard deviation of 0.43 μg/ml, and Cu concentrations increased with gestational age up to 20 weeks. The random effect meta-analysis across the 18 cohorts revealed that 1 μg/ml increase in maternal Cu concentration before the third trimester was associated with a higher risk of PTB with an OR of 1.30 (95% CI: 1.08 to 1.57) and shorter gestational duration of 1.64 days (95% CI: 0.56 to 2.73). The estimated effects were generally consistent across all sites. In the Malawi cohort, higher maternal Cu concentration, concentrations of multiple APRs and infections (malaria and HIV) were correlated and associated with greater risk of PTB and shorter gestational duration.

**Interpretation:** Our study supports a robust negative association between maternal mid-gestation Cu concentration and gestational duration and a positive association with risk for preterm birth. Cu concentration was strongly correlated with APRs and infection status suggesting its potential role in inflammation, a pathway implicated in the mechanisms of PTB. Therefore, maternal Cu could be used as a potential marker of the integrated inflammatory pathways during pregnancy and risk for preterm birth.

## INTRODUCTION

Preterm birth (PTB), defined as birth before 37 completed weeks (259 days) of gestation is the leading cause of perinatal morbidity and mortality worldwide (1). Globally, it is estimated that approximately 15 million babies are born preterm every year, with an average PTB rate of ∼11% (2), ranging from 5% to 18% with higher rates occurring in sub-Saharan African and South Asian low and middle-income countries. Despite the global burden the underlying drivers of PTB are uncertain. In particular, little is known about the role of maternal essential trace metals in contributing to or predicting PTB.

Copper (Cu) is an essential trace element regulating several critical biological processes through incorporation into copper-dependent proteins. These cuproproteins serve critical cellular homeostatic functions in maintaining redox status, antioxidant defenses and modulating inflammatory processes (3-7). Ceruloplasmin is the major Cu-carrying protein in the blood, carrying ∼ 75-95% of circulating Cu (8). The main function of ceruloplasmin is to oxidize ferrous iron (Fe^2+^) to the less damaging ferric iron (Fe^3+^) which enables the iron to be bound by transferrin, the major iron-transport protein. Cu in the form of ceruloplasmin possesses antioxidant activity by preventing free radical damage (8). Cu also has multiple actions implicated in the modulation of inflammation including the rise in ceruloplasmin (9, 10) whose expression levels increase during infection, stress, and inflammation. Ceruloplasmin is an acute phase reactant (APR) protein, the expression of which increases with systemic inflammation similar to C-reactive protein (CRP) and α1-acid glycoprotein (AGP) (11). Animal studies have demonstrated the role of Cu in modulation of inflammation and lipid peroxidation (10, 12-14). Spontaneous PTB is thought to be prompted by a cascade of inflammatory events, leading to cytokine upregulation and subsequent induction of uterine activity by promoting the expression and release of uterotonic factors (15). Maternal Cu concentration during pregnancy increase through early pregnancy and reportedly plateau by mid 2^nd^ trimester (16). Recently, some studies have suggested higher maternal or cord blood Cu levels were associated with increased risk of PTB (17, 18) while some others suggested an increased risk of PTB with Cu deficiency (19, 20).

In this study, we aimed to examine the association of maternal Cu concentration during early- and mid-pregnancy with PTB risk and gestational duration in a large number of samples collected from geographically diverse study cohorts with different social and ancestral backgrounds and varying degrees of environmental exposures. We leveraged the APRs and infection (malaria and HIV) data collected from pregnant women at the same gestational age as Cu concentration from the Malawi cohort to investigate the correlations between maternal Cu concentration with inflammation and infection.

## METHODS

### Study design and participants

The International Consortium on Selenium, Genetics and Preterm Birth is a Bill & Melinda Gates Foundation (BMGF) funded project to study the possible associations between maternal prenatal trace metals and the risk of PTB using data and samples from established diverse birth cohort worldwide. The consortium comprises 18 international pregnancy cohorts across a wide geographic distribution (**Figure 1**) with Cincinnati Children’s Hospital Medical Center (CCHMC) serving as the coordinating hub (21). Our study protocol was approved by the Institute Review Board (IRB) of the CCHMC and by the corresponding Ethics Committees of each participating institution. Among the participating sites, Malawi (iLiNS-DYAD) (22) and Bangladesh (MDIG) (23) cohorts were intervention trials and USA, CA (CPPOP) was a case-control study. All the other cohorts were designed to enroll eligible pregnant women in the community, or at hospitals. The description and study characteristics of these cohorts are provided in **Supplementary Text 1** and **Supplementary Table 1**.

**Figure 1.**
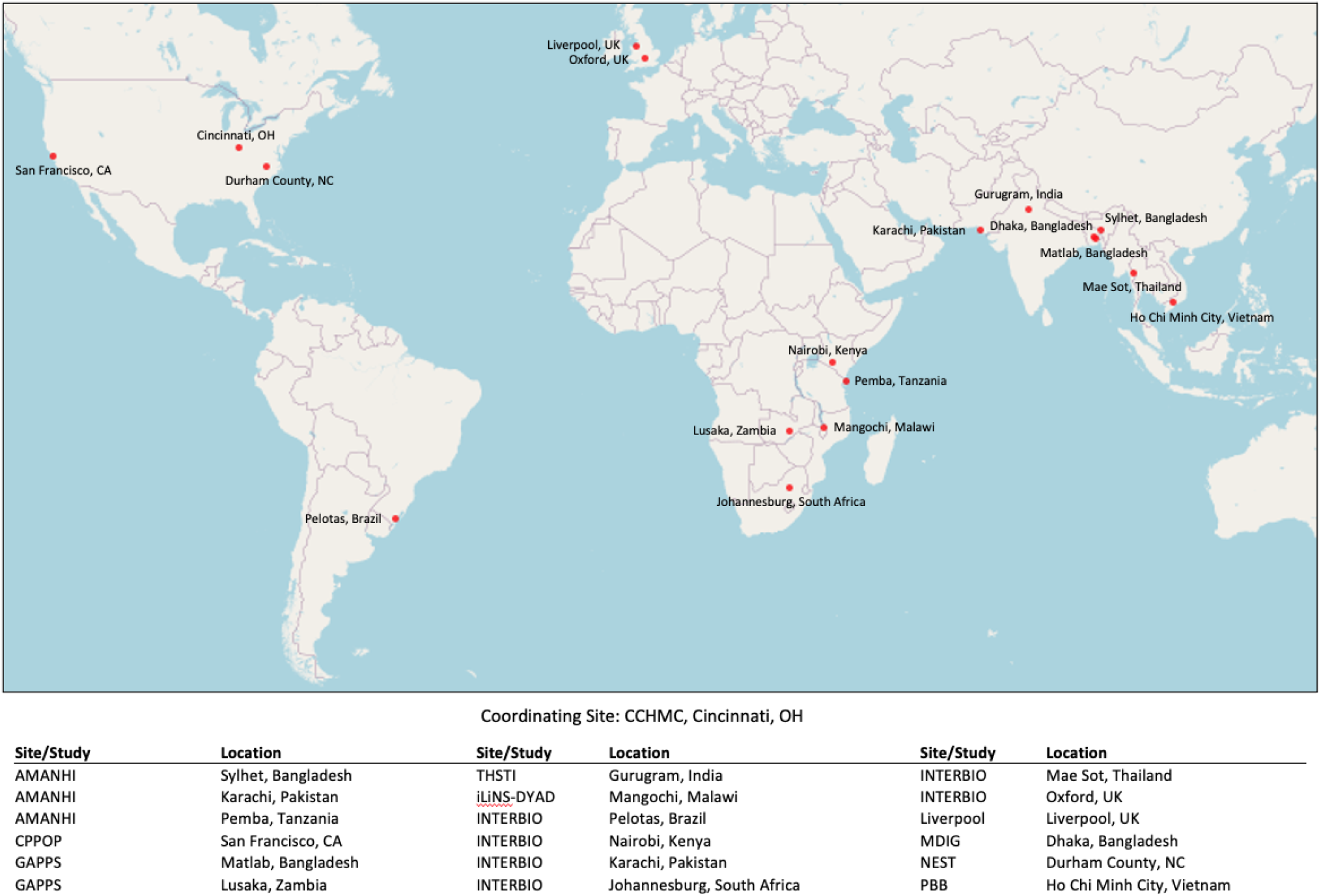
Geographic location of study sites.

### Samples and sampling data

Demographic, prenatal, delivery and fetal/newborn data (**Supplementary Table 2)** as collected by the individual sites according to their local protocols were shared with the coordinating hub (CCHMC). The data collected from Bangladesh (GAPPS), Bangladesh (MDIG) (23), Vietnam (PBB), USA (NEST; CPPOP) (24, 25), and all AMANHI cohorts (26) were case/control (preterm/term) samples. The data collected from other sites, including Malawi (iLiNS-DYAD) (22), Zambia (GAPPS), India (THSTI)(27) and the six INTERBIO-21^st^ sites (28) were population or hospital based samples. Gestational age dating was assigned at the site level by ultrasound, last menstrual period (LMP) or both (**Supplementary Table 1**). Preterm cases were defined as birth prior to 37 weeks of gestation and term controls as birth at 37 weeks or later. We excluded stillbirths and multi-gestational pregnancies.

### Copper measurement

Cu concentrations were measured in maternal plasma or serum (29) stored at -70 □ or -80 □ freezers before and after use at the CCHMC Biobank (**Supplementary Table 1**). To mitigate the potential batch effect, samples from each site were randomized prior to analysis in batches. Inductively coupled plasma mass spectrometry (ICP-MS) measurements of Cu concentrations were performed using Agilent 7700 ICP-MS (Agilent Technologies) at the laboratory of Clinical Chemistry and Biochemistry, the University of Cincinnati as described in detail in the protocol (**Supplementary Text 2**). The samples from India (THSTI) were analyzed at the Inter University Accelerator center at National Geochronology Facility (IUAC, Delhi, India) using the same method and protocol as CCHMC. The samples from Bangladesh (MDIG) were analyzed at the Centers for Disease Control and Prevention (CDC, Atlanta, GA).

### Acute Phase Reactants in Malawi cohort

We obtained plasma concentrations of three APRs (C-reactive protein (CRP), α1-acid glycoprotein (AGP) and albumin (ALB) measured in Malawi (iLiNS-DYAD) cohort samples collected at the same time for Cu measurement. Plasma concentrations of CRP and AGP were measured by immunoassay using a COBAS Integra Analyzer (Roche Diagnostics, Mannheim, Germany). All samples were analyzed singularly, except for 5%, which were randomly selected to be analyzed in duplicate. None of the samples analyzed in duplicate had a coefficient of variation (CV) greater than 5%. HIV infection at study enrollment was tested with a whole-blood antibody rapid test (Alere Determine HIV-1/2; Alere Medical). Malaria during pregnancy was diagnosed on-site from finger-prick blood samples using the rapid diagnostic test Clearview Malaria Combo (British Biocell International).

### Statistical analysis

Phenotypic data from participating study sites were harmonized by applying a uniform data structure and consistent coding rules for key variables (e.g, gestational duration, maternal age, maternal height, and fetal sex). The distributions of gestational duration and Cu concentrations for each site were visually inspected using histograms and violin plots. Outliers for the gestational duration and Cu concentrations were detected based on fitting with appropriate probability distributions and removed from further association analysis.

To determine the covariates to be included in the association analysis, we first examined the correlation of PTB and gestational duration with other covariates as well as that between Cu concentration and other covariates at each site using Pearson correlation. The DerSimonian-Laird (DSL) random-effect meta-analysis was used to combine the correlation coefficients across the study sites. Variables significantly correlated (p < 0·05) with either PTB or gestational duration or Cu concentration were included as covariates. For each site, we estimated the association between maternal Cu concentration and PTB (and gestational duration as a continuous variable) using logistic (for PTB) or linear (for gestational duration) regression analysis with selected covariates. Random-effect meta-analysis was used to combine the results from different cohorts and between-study heterogeneity was checked using Cochran’s Q test.

Some of the cohorts used case/control samples (**Supplementary Table 1 and 3**) with different case/control ratios. Because regression analysis of gestational duration as a continuous variable in non-random samples could potentially introduce bias in effect size estimation, we conducted regression analysis weighted by the inverse of sampling probability (IPW) based on their case/control status (**Supplementary Table 3**).

All analyses were performed with Microsoft R Open 4.0.2. The cross-site meta-analysis of associations and correlations were conducted using metafor and metocor packages.

## RESULTS

### Gestational duration, PTB and their correlations with other covariates

Pregnancy phenotype and birth outcomes of 11,160 pregnancies were obtained from 18 study sites (**Supplementary Table 1**). Among these, 10,449 singleton live births had a gestational age estimated in days (gday) and maternal plasma or serum Cu concentrations measured (**Figure 2**). The characteristics of these mothers (e.g., age, height and gestational duration are summarized by site (**Table 1**). After removing three outliers, the gestational duration followed a Weibull distribution with a mean of 268 days and a median of 273 days ranging from 147 to 312 days (distribution parameters: shape: 21.1, scale: 276.0) (**Supplementary Figure 1**). The distributions of gestational days in term (gday ≥ 259 days) and preterm (gday < 259 days) births from each site are shown in **Supplementary Figure 2**.

**Figure 2.**
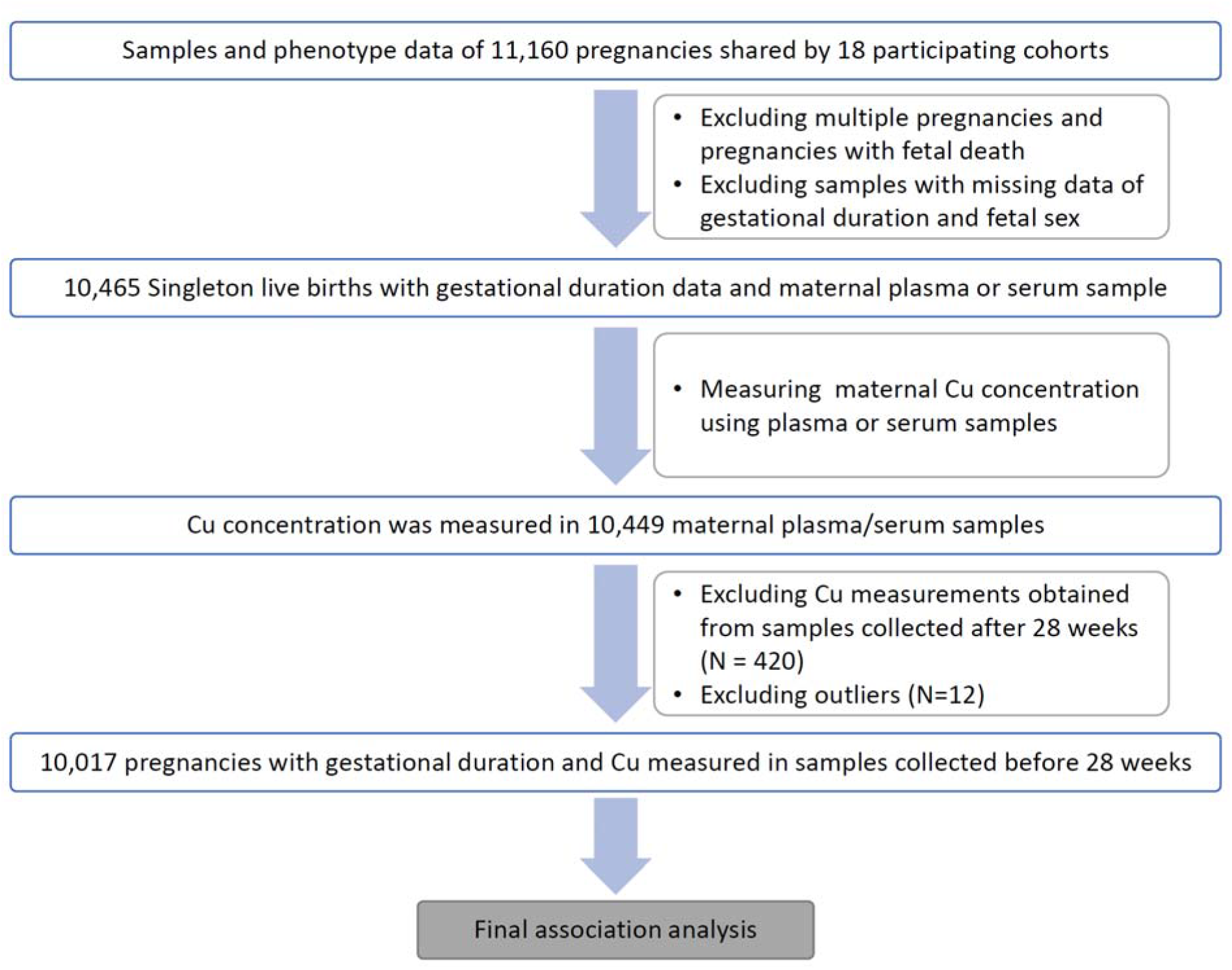
Flow chart of the study illustrating the total number of subjects, inclusion/exclusion criteria. *(N=420) excluded from final association analysis: 416 with samples collected at gestational age >28wks and 4 samples with missing information. *(N=12) outliers excluded: 3 for gestational duration; 9 for Cu concentration

**Table 1.** Demographic characteristics of study subjects.

| Site | Sample size | Preterm |  | Sex |  | Maternal Age |  | Maternal Height |  | GA at delivery |  | GA at sampling |  | Birth Weight |  |
| --- | --- | --- | --- | --- | --- | --- | --- | --- | --- | --- | --- | --- | --- | --- | --- |
|  |  | Term | Preterm | Male | Female | Mean | SD | Mean | SD | Mean | SD | Mean | SD | Mean | SD |
| Bangladesh (AMANHI) | 506 | 253 (50.0%) | 253 (50.0%) | 239 (47.2%) | 267 (52.8%) | 23.6 | 4.5 | 149.2 | 5.6 | 260.7 | 20 | 95.8 | 23.1 | 2516.1 | 495.2 |
| Bangladesh (GAPPS) | 258 | 172 (66.7%) | 86 (33.3%) | 132 (51.2%) | 126 (48.8%) | 23.9 | 5.9 | 151.8 | 5.5 | 267.8 | 18.7 | 158.8 | 4.7 | 2722.7 | 581.1 |
| Bangladesh (MDIG) | 205 | 135 (65.9%) | 70 (34.1%) | 105 (51.2%) | 100 (48.8%) | 23.4 | 4.5 | 151.4 | 5.6 | 265.7 | 14.3 | 143.9 | 13.5 | 2664.6 | 378.4 |
| Brazil (INTERBIO) | 389 | 344 (88.4%) | 45 (11.6%) | 212 (54.5%) | 177 (45.5%) | 28.5 | 5.4 | 162.5 | 6.4 | 270.4 | 10.8 | 132.2 | 53.9 | 3147.1 | 464.3 |
| India (THSTI) | 506 | 435 (86.0%) | 71 (14.0%) | 281 (55.5%) | 225 (44.5%) | 23.4 | 3.8 | 153.2 | 6 | 271.1 | 14.4 | 91.6 | 25.1 | 2744.8 | 492.2 |
| Kenya (INTERBIO) | 553 | 528 (95.5%) | 25 (4.5%) | 293 (53.0%) | 260 (47.0%) | 30.4 | 4.1 | 161.8 | 5.8 | 278.3 | 11.1 | 112.5 | 40.6 | 3267 | 463.8 |
| Malawi (iLiNS-DYAD) | 1212 | 1120 (92.4%) | 92 (7.6%) | 587 (48.4%) | 625 (51.6%) | 25.2 | 6.2 | 156.1 | 5.7 | 276 | 14.3 | 117.7 | 14.9 | 2976.6 | 449.5 |
| Pakistan (AMANHI) | 348 | 233 (67.0%) | 115 (33.0%) | 189 (54.3%) | 159 (45.7%) | 26.3 | 5.1 | 154.8 | 6.1 | 265.5 | 16.5 | 95.1 | 24.6 | 2684.4 | 500.1 |
| Pakistan (INTERBIO) | 516 | 413 (80.0%) | 103 (20.0%) | 251 (48.6%) | 265 (51.4%) | 30.1 | 4.6 | 158 | 5.9 | 264.9 | 13.5 | 103.5 | 35.6 | 2876.5 | 480.7 |
| South Africa (INTERBIO) | 352 | 299 (84.9%) | 53 (15.1%) | 181 (51.4%) | 171 (48.6%) | 30.2 | 5.8 | 159 | 6.9 | 269.4 | 17.5 | 88.2 | 19.6 | 2940.3 | 588.8 |
| Tanzania (AMANHI) | 351 | 234 (66.7%) | 117 (33.3%) | 174 (49.6%) | 177 (50.4%) | 27.9 | 6.6 | 155.2 | 5.9 | 267.5 | 19.6 | 99.1 | 23.1 | 3111.9 | 592.5 |
| Thailand (INTERBIO) | 514 | 485 (94.4%) | 29 (5.6%) | 266 (51.8%) | 248 (48.2%) | 26.2 | 6.1 | 151.8 | 5.1 | 275.6 | 11.5 | 114.4 | 37.1 | 2965.8 | 457.3 |
| UK (INTERBIO) | 648 | 594 (91.7%) | 54 (8.3%) | 342 (52.8%) | 306 (47.2%) | 31.1 | 4.8 | 165.3 | 6.5 | 275.9 | 14.6 | 89.9 | 20.3 | 3301.1 | 586 |
| UK (Liverpool) | 525 | 424 (80.8%) | 101 (19.2%) | 271 (51.6%) | 254 (48.4%) | 30.6 | 4.9 | 164.8 | 6.3 | 267 | 21.7 | 140.8 | 9.5 | 3141.2 | 730.3 |
| USA, California (CPPOP) | 966 | 484 (50.1%) | 482 (49.9%) | 505 (52.3%) | 461 (47.7%) | 30 | 6.1 | 161.6 | 7.3 | 249.8 | 29.9 | 115.7 | 7.7 | 2763.1 | 923 |
| USA, North Carolina (NEST) | 657 | 438 (66.7%) | 219 (33.3%) | 363 (55.3%) | 294 (44.7%) | 28.3 | 6.1 | 162.8 | 7.7 | 263.1 | 22.5 | 161.1 | 90.5 | 2999.6 | 750.4 |
| Vietnam (PBB) | 970 | 651 (67.1%) | 319 (32.9%) | 495 (51.0%) | 475 (49.0%) | 29.1 | 4.6 | 156 | 4.8 | 264.4 | 18.9 | 149 | 6.2 | 2959.9 | 613.5 |
| Zambia (GAPPS) | 973 | 853 (87.7%) | 120 (12.3%) | 478 (49.1%) | 495 (50.9%) | 27.7 | 5.8 | 160.4 | 6.5 | 271.6 | 18.2 | 137.3 | 27.2 | 3008.7 | 591.7 |
| TOTAL | 10449 | 8095 (77.5%) | 2354 (22.5%) | 5364 (51.3%) | 5085 (48.7%) | 27.8 | 5.9 | 158.2 | 7.6 | 267.9 | 19.9 | 120.6 | 39.8 | 2956.5 | 632.9 |

We examined the correlation of PTB and gestational duration with other covariates (maternal age, height, fetal sex, and gestational age at sampling) in each participant site (**Supplementary Figure 3**). Meta-analysis using the DSL method showed that PTB risk was significantly correlated with maternal height and fetal sex. Similarly, gestational duration was also significantly correlated with maternal height (shorter mothers had shorter gestational duration) and fetal sex (males had shorter gestational duration).

### Maternal prenatal Cu concentration and its correlations with other covariates

Cu concentrations were successfully measured in 10,449 mothers. After excluding twelve outliers, the Cu concentrations followed a normal distribution (**Supplementary Figure 4**) with a mean of 1.92 μg/ml and standard deviation of 0.43 μg/ml. The Cu concentrations varied across different sites (**Supplementary Figure 5, Supplementary Figure 6)** and across different experimental batches for each site (**Supplementary Figure 7**). The highest average Cu was observed in the Bangladesh (MDIG) cohort with a mean level of 2.2 μg/ml, and the lowest Cu was in the Bangladesh (AMANHI) cohort with a mean concentration of 1.4 μg/ml (**Table 2**). The Cu concentrations increased with gestational age (see below); however, even after adjustment for gestational age at sampling the Cu concentrations still showed between-site differences (**Supplementary Figure 5B**).

**Table 2.** Summary statistics of maternal Copper concentration in different study sites.

| Site | N | Mean | SD | Median | Min. | Max. |
| --- | --- | --- | --- | --- | --- | --- |
| Bangladesh (AMANHI) | 506 | 1.36 | 0.34 | 1.34 | 0.62 | 2.34 |
| Bangladesh (GAPPS) | 258 | 1.85 | 0.32 | 1.82 | 0.97 | 3.22 |
| Bangladesh (MDIG) | 205 | 2.21 | 0.4 | 2.18 | 1.06 | 3.48 |
| Brazil (INTERBIO) | 389 | 1.81 | 0.34 | 1.8 | 0.84 | 3.45 |
| India (THSTI) | 504 | 1.7 | 0.48 | 1.67 | 0.16 | 3.74 |
| Kenya (INTERBIO) | 553 | 1.94 | 0.37 | 1.92 | 0.99 | 3.63 |
| Malawi (iLiNS-DYAD) | 1211 | 2.01 | 0.36 | 1.99 | 0.93 | 3.42 |
| Pakistan (AMANHI) | 346 | 2.01 | 0.5 | 1.95 | 0.96 | 3.72 |
| Pakistan (INTERBIO) | 516 | 1.95 | 0.45 | 1.93 | 0.84 | 3.83 |
| South Africa (INTERBIO) | 352 | 1.99 | 0.41 | 1.96 | 0.82 | 3.44 |
| Tanzania (AMANHI) | 350 | 1.88 | 0.4 | 1.87 | 0.9 | 3.16 |
| Thailand (INTERBIO) | 514 | 1.7 | 0.36 | 1.7 | 0.63 | 3.55 |
| UK (INTERBIO) | 644 | 1.74 | 0.38 | 1.71 | 0.69 | 3.17 |
| UK (Liverpool) | 525 | 2.1 | 0.41 | 2.06 | 0.23 | 3.57 |
| USA, California (CPPOP) | 966 | 2.08 | 0.37 | 2.07 | 0.14 | 3.57 |
| USA, North Carolina (NEST) | 657 | 1.98 | 0.49 | 1.96 | 0.23 | 3.65 |
| Vietnam (PBB) | 969 | 1.98 | 0.39 | 1.94 | 1.04 | 3.8 |
| Zambia (GAPPS) | 972 | 2.05 | 0.35 | 2.02 | 1.11 | 3.75 |
| TOTAL | 10437* | 1.92 | 0.43 | 1.91 | 0.14 | 3.83 |

\*Total number of samples with maternal Cu concentrations measured was 10449. Total of 12 outliers excluded: 3 for gestational duration and 9 for Cu concentration

We examined the correlation of maternal Cu concentration with other covariates in each site. When combined across sites, the Cu concentration across sites was significantly positively correlated with maternal age (ρ = 0.05, p = 0.0017), negatively correlated with maternal height (ρ = -0·05, p = 0.0001) and was higher in mothers with female babies (p = 0.01). Cu concentration was also positively correlated with selenium concentration measured from the same samples (ρ = 0.14, p = 0.0014) (Se data was not available in THSTI samples). In addition, there was a strong positive correlation between Cu concentration and gestational age at the time of sample collection (ρ = 0.28, p = 3.3e-9) (**Supplementary Figure 8**).

### Change of maternal prenatal Cu concentration during pregnancy

To inspect the change of maternal Cu concentration visually across different gestational ages, we generated boxplots of Cu concentration at each week of gestational age of sampling for the 10,432 samples collected between 5 weeks to 41 weeks (**Figure 3**). The mean maternal Cu concentration increased substantially during early gestation, from a mean at 1.3 μg/ml to 2.0 μg/ml between week 5 to week 16 and plateaued and gradually reached 2.2 μg/ml near delivery with some fluctuations. Given the non-linear relationship between maternal Cu and gestational age at sampling, the first two polynomials of gestational age of sampling were included in later regression analysis.

**Figure 3.**
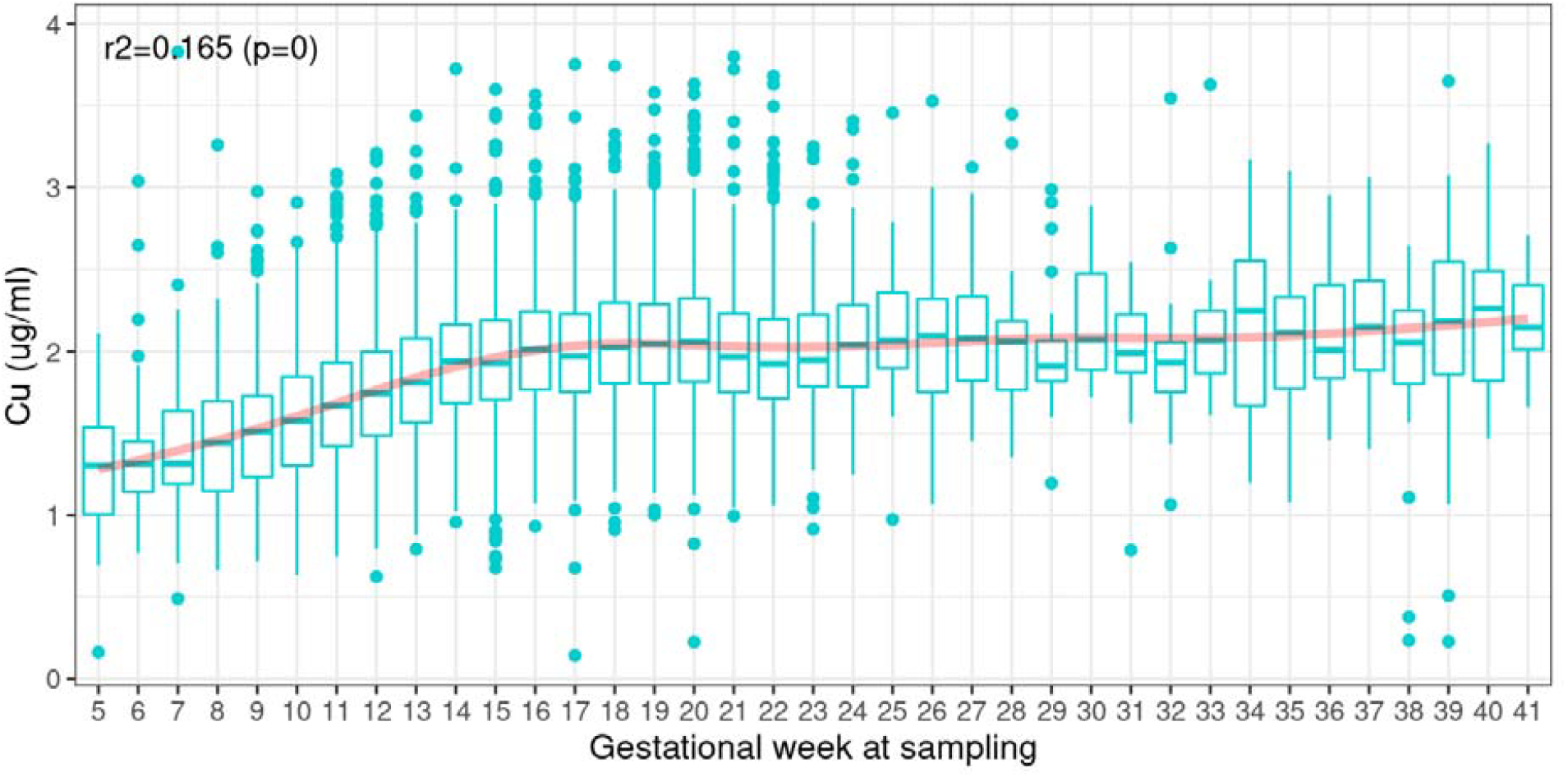
Copper concentration at each gestational week of sampling. The red shaded curve shows the LOESS smoothing of Cu concentration at different gestational weeks.

The gestational age at sample collection varied substantially from site to site and at some sites, there were some samples collected after the second trimester (≥ 28weeks of gestation) (**Supplementary Figure 9**). To minimize the bias introduced by these samples (e.g., exclusion of extremely PTB and the non-linear increase of maternal Cu concentration), we excluded 420 samples that were collected at 28weeks of gestation or later (plus four samples without a known date of sample collection) from the final association analysis.

### Association of maternal Cu concentration with PTB and gestational duration

We examined the association of maternal Cu concentration before the third trimester with gestational age at sample collection < 28 weeks with PTB and gestational duration in each individual site and then combined the results using meta-analysis (**Figure 4**). In total, the associations were tested in 10,017 pregnancies (**Figure 2**). The covariate factors that found to be significantly associated (p < 0·05) with either gestational duration or Cu concentration were incorporated as covariates. These included maternal age (mage), maternal height (ht), fetal sex (fsex) experimental batch (batch) and the first two polynomials of gestational days at sample collection [gday(sample)]. Given the enrichment of preterm cases in the case–control studies that could potentially introduce bias, we conducted the IPW analysis in the eight case/control data sets in the regression analysis of gestational duration to correct for the sampling bias (**Figure 4B, Supplementary Table 3**).

**Figure 4.**
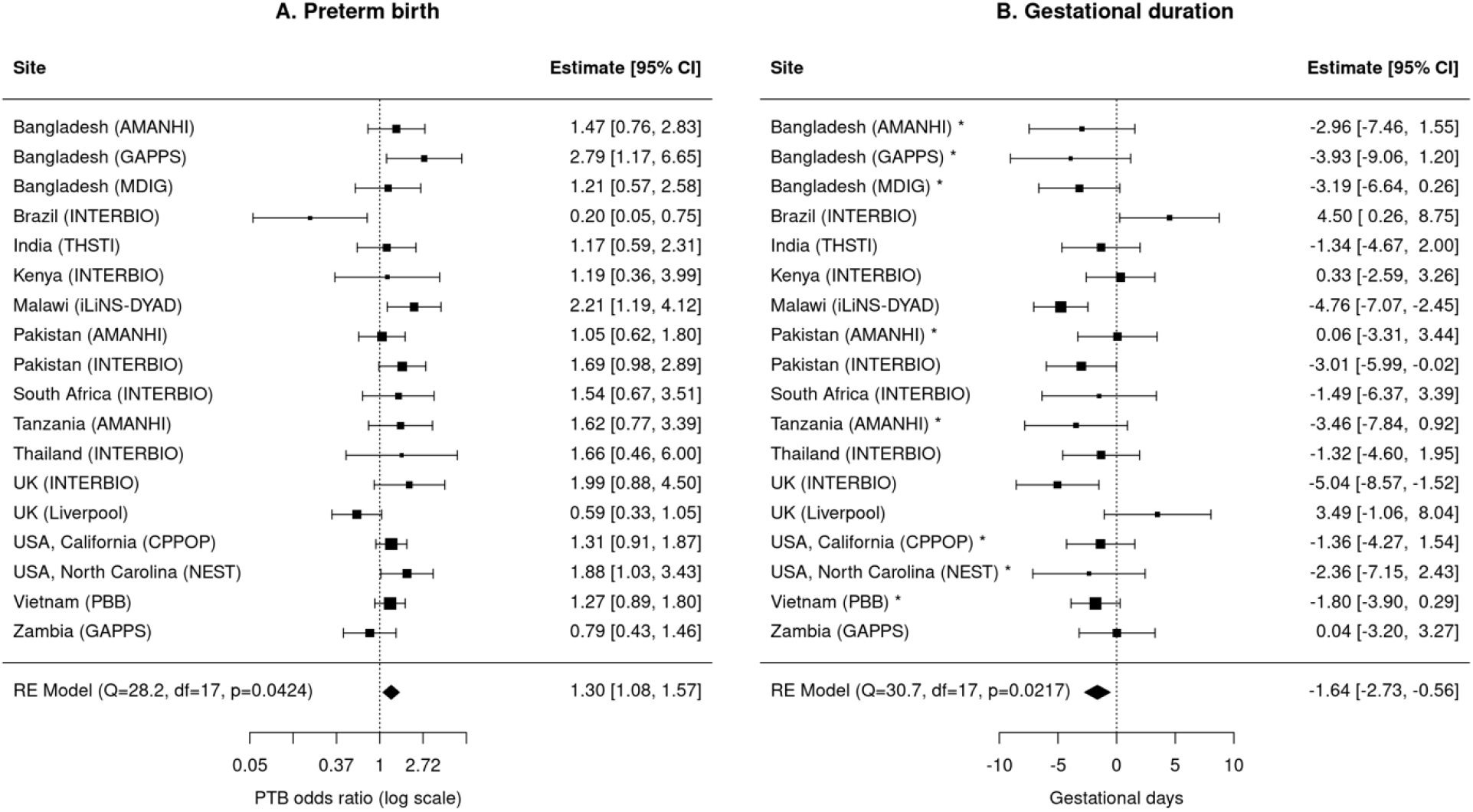
Meta-analysis of the association of maternal Copper concentration with PTB (A) and gestational duration (B) * Sites with case/control sampling were corrected by IPW in regression analysis of gestational duration (B).

In the combined meta-analysis and in several of the individual cohorts, higher maternal Cu concentration was associated with higher risk of PTB and shorter gestational duration (**Figure 4**). There was some between-cohort heterogeneity, for example, the observed associations in Brazil (INTERBIO) and the UK (Liverpool) pointed in the opposite direction. Pooled effect size estimates were an OR= 1.30 (95% CI: 1.08 to 1.57) for PTB or 1.64 days (95% CI: 0.56 to 2.73) shorter gestation per 1 μg/ml increase in Cu concentration. The association between gestational age at sampling adjusted Cu concentration and gestational duration followed a linear relationship (**Supplementary Figure 10**) within a wide range of Cu concentration (up to ±3sd). The fraction of PTB cases also showed a monotonic increase from the lowest to the top quartile group of the adjusted Cu concentration (**Supplementary Figure 11**).

### Correlation of maternal Cu concentration with acute phase reactants and infections in the Malawi cohort

In the Malawi (iLiNS-DYAD) cohort, we examined correlations between maternal Cu concentration and common analytes including acute phase reactants (CRP, AGP and ALB), and infections (HIV and malaria) in 1239 samples collected at enrollment. Participants of the Malawi cohort were enrolled from four health facilities that covered mostly one continuous area near Lake Malawi (21). Some of the common analytes (including CRP, AGP) followed log-normal distributions and were, therefore, log-transformed in the statistical analyses. As significant differences in gestational duration, gestational days at sampling, maternal Cu concentration and many analytes were observed among these four Malawi subsites (**Supplementary Figure 12**), we performed statistical analyses stratified by these four subsites using similar methods as we did in the meta-analyses across the 18 major study sites.

Similar to the maternal Cu concentration (**Figure 3**), the concentrations of all the analytes were influenced by gestational age at sampling (**Supplementary Figure 13**). Therefore, we calculated adjusted values of these variables using the first two polynomials of gestational age at sampling and tested their pairwise correlations and their associations with gestational duration and PTB. Strong pairwise correlations were observed among maternal Cu and almost all the APRs, malaria, and HIV infections (**Figure 5**). These measurements were also significantly correlated with gestational duration or PTB risk. Among these, higher maternal Cu, CRP, and AGP were associated with a higher risk of PTB and shorter gestational duration, and higher ALB was correlated with reduced risk of PTB and longer gestational duration. The infection rates of HIV and malaria were also positively and negatively correlated with maternal age at pregnancy respectively.

**Figure 5.**
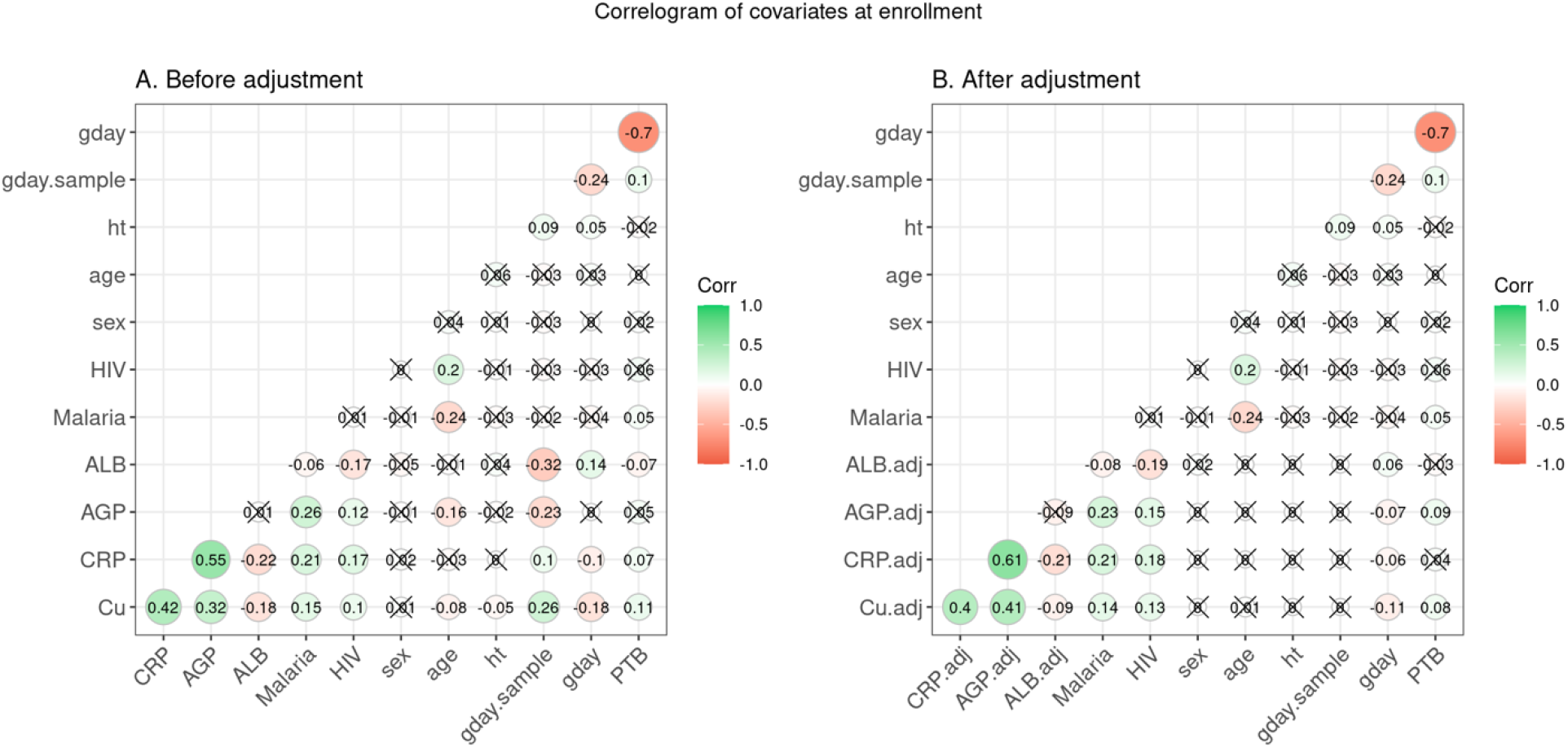
Correlogram of Cu concentration, APRs, and phenotype measured in Malawi (iLiNS-DYAD) cohort. Red shading indicates a negative correlation and green a positive correlation, with intensity reflecting the magnitude of the correlation. Correlations with p-value > 0.05 were indicated by crosses. A. Pairwise correlation before adjustment for gestational age at sampling. B. Pairwise correlation after adjustment for gestational age at sampling.

The magnitudes of some of the pairwise correlations changed after adjustment for gestational age at sampling (**Figure 5**). Particularly, the correlations of AGP with PTB or gestational duration were only significant after adjustment, and the correlations of ALB with PTB or gestational duration were less significant after adjustment. After adjustment for gestational age at sampling the magnitude of the observed association between Cu and AGP increased by 27%, and the magnitude of the association of Cu with ALB reduced by 50% (**Supplementary Figure 14)**. Because Cu and the APRs were correlated with each other and all of them were correlated with PTB or gestational duration at different magnitudes (**Supplementary Figure 15**), we examined whether the observed associations of Cu with PTB and gestational duration were modulated by APRs, or HIV and malaria infections. The estimated associations of Cu with PTB and gestational duration were attenuated after the inclusion of other analytes or infections as a covariate, and this was most apparent when adjusting for CRP or AGP (**Supplementary Figure 15A**). If including all the APRs and infections as covariates (ALL), the effect sizes of Cu reduced substantially; however, the residual association between Cu and gestational duration was still marginally significant (**Supplementary Figure 15B**).

## DISCUSSION

In this meta-analysis of diverse cohorts from multiple low and middle income Asian and sub-Saharan African countries as well as in high-income countries (UK and US), we measured maternal prenatal circulating Cu concentrations and tested their association with the risk of PTB and gestational duration. Our results demonstrate that maternal Cu concentrations were normally distributed and there was substantial variation among different sites. The lowest Cu concentration was observed in Bangladesh (AMANHI) samples, which was partially due to the early gestational age of sampling at this site (<20 weeks). However, even after adjusting for gestational age at sample collection, important differences remained. Maternal Cu levels even differed among sites in geographic and cultural proximity (e.g., three Bangladesh sites), indicating that local dietary and environmental factors influence Cu concentrations. We also observed that Cu concentrations were higher in older women, women with shorter stature, and among women gravid with female fetuses. Maternal Cu concentrations increased substantially during early pregnancy – the mean increased by ∼50% from week 5 to 16 and then reached a plateau around 2.0μg/ml.

For the association analysis of maternal Cu concentrations (before the third trimester) with PTB and gestational duration, we found significant positive associations between maternal Cu and PTB risk and negative associations with gestational duration. The overall estimated effect sizes are OR of 1.30 for PTB risk and 1.64 days shorter gestation per 1 μg/ml increase in maternal Cu concentration. Although among-site heterogeneity was observed, the estimated effects were generally consistent across sites, except for Brazil (INTERBIO) and UK (Liverpool), in which the associations point in the opposite directions. Overall, the results of our meta-analysis agree with previous studies which also showed higher maternal Cu levels are associated with increased risk of PTB (18, 30). In addition, we demonstrated that the association between maternal Cu concentration and gestational duration followed a linear relationship and the PTB rates increased monotonically in mothers with higher quartiles of Cu.

In the Malawi samples (12% of the total samples), higher maternal Cu, CRP, and AGP were associated with a higher risk of PTB and shorter gestational duration, and higher ALB was correlated with reduced risk of PTB and longer gestational duration. These results indicate that Cu could be used as an indicator to capture the associations between acute phase proteins (e.g., CRP and AGP) and gestational duration.

### Significance and implications of this study

Our findings contribute to an emerging literature focused on the association of Cu status and pregnancy outcomes, especially the risk of PTB and gestational duration (17, 18, 30, 31). Cu is an essential trace element involved in numerous biological processes, and disorders of Cu metabolism in pregnancy, either deficiencies or excesses, can lead to adverse pregnancy outcomes such as preeclampsia and PTB (15). However, the effect of Cu status on the risk of PTB is not well understood with some suggesting increased PTB and others reporting contradicting results (19, 20). A recent case-control study of pregnant women from Malawi showed that higher maternal Cu at delivery was associated with increased risk of PTB (30). Hao et al. collected plasma and serum at the first antenatal visit between 4 and 22 of gestation and found that the overall median maternal serum Cu concentrations were significantly higher for preterm births than for term births in the Chinese population (18). However, there are also reports on gestational length that found contradicting results. Evidence supporting the potential involvement of Cu in PTB risk includes a study of the Maan’Shaan Birth cohort in China, which showed that relatively low umbilical cord Cu levels were associated with higher risk of PTB and early-term birth (19). These discrepancies may be due to differences in population, study design, maternal or fetal origin of samples or timing of sampling in pregnancy. Most previous studies have been either case-control studies or mostly focused on a single geographic region and are generally based on small sample sizes.

The present study is the most extensive investigation of the association between early and mid-pregnancy Cu concentration and gestational duration and PTB in global populations, including cohorts from low-income Asian and sub-Saharan African countries with a very high baseline PTB risk. Overall, our results support a consistent association between maternal Cu concentration and PTB and gestational duration. We cannot draw causal inference from this observational study. It is possible that Cu may play some functional roles in inflammation and antioxidant mechanisms, the pathways that were implicated in the mechanisms of PTB. Without ceruloplasmin measurements at the same gestational age as Cu measurements, we cannot rule out the confounding effects of the inflammation leading to upregulation of ceruloplasmin which in turn drives up Copper. However, even after adjusting for various other inflammatory markers, our results still showed that Cu significantly associated with gestational duration suggesting maternal Cu during pregnancy could be a marker of the integrated inflammatory pathways implicated in preterm birth.

The mechanisms underlying the association between higher serum or plasma Cu and PTB are still not fully understood. We propose that maternal circulating Cu concentrations at early or mid-gestation reflect heightened inflammation in those pregnancies destined for deliver preterm spontaneously. The hierarchy of biological activities of Cu calls for biomarkers informative at different levels of Cu exposure assessing Cu intake, placental or tissue Cu, Cu excretion and Cu biological function. Plasma or serum Cu concentration provides valuable information about the Cu status over a wide range of Cu intake; however, there is need for additional information regarding Cu particularly for assessing the Cu status and its specific mechanistic role in those high risk for PTB. Epidemiological reports and research examining the effects of Cu along with specific markers of inflammation and oxidation such as ceruloplasmin which is Cu-dependent protein at the same gestational age collected using a standardized protocol are required.

### Strength and limitations

Samples and phenotypic data were retrieved from existing biorepositories collected several years ago in different studies. Although we harmonized and analyzed a set of key variables known to be associated with PTB and gestational duration, we were unable to include some important environmental and socioeconomic factors in the analysis due to missing or incomplete data. We excluded stillbirth due to missing data on cause-of-death, underreporting, and lack of comparability in reporting stillbirths, especially in low- and middle-income countries regarding the birth weight and gestational age criteria. Also, our study only focused on overall PTB and haven’t separated iatrogenic from SPB as the data is missing from several of the cohorts. This is a very significant confounder that was not controlled for, and likely had an impact on the statistical significance and strength of the association found across the cohorts. It will be key to include these variables across cohorts in future studies.

There were differences in how gestational age was determined and distributed across cohorts. Some cohorts determined the duration by ultrasound fetal biometry (parameters varied across cohorts) whereas others used LMP (or both). This different dating methodology between studies may have introduced some noise into the analysis. PTB rates reported in some low- and middle-income cohort studies appear to be low, and this might be due to underreporting and geographic location of the recruitment site. Also, some cohorts were enriched for PTB samples, and the distribution of gestational duration did not follow a normal distribution. Although regression analysis is generally robust regardless of meeting the normality assumption and we utilized IPW to adjust for the case/control sampling in regression analysis of gestational duration, this difference in study design and data collection may have introduced some bias in these analyses.

Also, of note with respect to the study limitations is that there was large variation in gestational age when the plasma/serum samples were collected. Given the gestational age at sample collection significantly correlated with the Cu concentration, we accounted for this variance by including only women with samples collected during the 1^st^ or 2^nd^ trimesters and adjusted for gestational age at sampling in the association analyses. Despite these methodological adaptations, it is possible that we may not have entirely accounted for the influence of gestational age at sample collection. More standardization with respect to the timing of sample collection and storage times may simplify these types of analyses in future studies.

Finally, although we tested several acute-phase proteins (e.g., CRP, AGP, ALB) in Malawi samples, these data were not available in all the study cohorts. Furthermore, we did not measure ceruloplasmin in this current study, which is presumably the most important acute-phase protein that influences the Cu concentration in blood. Future studies of maternal ceruloplasmin and Cu concentration will be essential to elucidate their role in pregnancy.

## Conclusions

Across 18 international birth cohorts with diverse ethnic backgrounds and geographic distribution, there were significant associations between maternal Cu concentration and PTB and gestational duration. These associations were consistent across most study sites and the association was monotonic and linear across the full range of Cu concentrations. Maternal Cu was strongly correlated with CRP, AGP, HIV and malaria infections measured in the Malawi samples. Adjustments for these APRs and infections attenuated the observed associations of maternal Cu with PTB and gestational duration, but the associations persist suggesting that maternal Cu concentration is an indicator of diverse factors that reflect the acute phase reaction and inflammation that ultimately impact the duration of pregnancy.

## Data Availability

All data produced in the present study are available upon reasonable request to the authors

## DECLARATIONS

### Ethical Approval and Consent to Participate

The study was approved by the Institute Review Board (IRB) of the Cincinnati Children’s Hospital Medical Center (URB 2017-3573) and by the corresponding Ethics Committees of each participating institution.

### Consent for publication

### Availability of Data and Materials

Data, the statistical code, and technical processes are available from the corresponding author on reasonable request.

### Funding

This work was supported by the Bill & Melinda Gates Foundation. The funders did not have any role in study design, data analysis, data interpretation, writing of the report, or submission for publication. The findings and conclusions of this article are solely the responsibility of the authors Bill & Melinda Gates Foundation (OPP1175128, OPP1152451)

Funders: Cohort Investigations

California Prediction of Poor Outcomes of Pregnancy (CPPOP): UCSF California Preterm Birth Initiative NEST: National Institute of Environmental Health Sciences (R21ES014947, R01ES016772, P30ES025128, and P01ES022831), the US Environmental Protection Agency (RD-83543701), the National Institute of Diabetes and Digestive and Kidney Diseases (R01DK085173), and the Duke Cancer Institute.

iLiNS-DYAD-M trial is funded by a grant to the University of California, Davis from the Bill & Melinda Gates Foundation [OPP49817]. A grant to the University of California, Davis from the Office of Health, Infectious Diseases, and Nutrition, Bureau for Global Health, U.S. Agency for International Development (USAID) under terms of Cooperative Agreement No. AID-OAA-A-12-00005 through the Food and Nutrition Technical Assistance III Project (FANTA), managed by FHI 360.

The Global Alliance to Prevention of Prematurity and Stillbirth (GAPPS) Biorepository Program funded by the Preventing Preterm Birth Initiative grant from the Bill & Melinda Gates Foundation [OPP1033514].

MDIG trial funded by The Bill & Melinda Gates Foundation [OPP1066764]

GARBH – Ini cohort funded by Department of Biotechnology (DBT), Government of India (BT/PR9983/MED/97/194/2013); and Bill & Melinda Gates Foundation (BMGF) (OPP1179761) through Grand Challenges India -Biotechnology Industry Research Assistance Council (GCI -BIRAC) Platform

### Authors’ Contributions

NM, HX and GZ prepared the first draft of the manuscript. LJM conceptualized and acquired the funding support for the study. The study was designed by NM, EB, JAL, GFC, JC, GZ and LJM. EB, JAL and NM conducted the ICP-MS analysis of the samples. HX, EB, NM and GZ compiled the data sets. HX and GZ developed the analytical pipeline and did the statistical analysis. JC coordinated all study related operations. All co-authors have contributed essential intellectual input, revisions of the manuscript, discussed the results, contributed to the revisions of the final manuscript, and approved the final version of the manuscript. The corresponding authors, NM and GZ had full access to the data and had final responsibility for the decision to submit for publication.

## Acknowledgements

We are grateful to all participating families and study personnel from 18 international pregnancy cohorts, who took part in this study. Payment for access to data and article-processing charges for this publication was covered by The Bill & Melinda Gates Foundation (Grant no: OPP1175128, OPP1152451). The authors would like to acknowledge the March of Dimes Prematurity Research Center Ohio Collaborative for their support to the original GWAS study that identified the EEFSEC gene and led to this study. The Aga Khan University would like to acknowledge Dr Yoshida Sachiyo and Dr Alexander Manu from the World Health Organization, community health workers, pregnant women and their families. The NEST study acknowledges the support from National Institute of Environmental Health Sciences, the US Environmental Protection Agency, the National Institute of Diabetes and Digestive and Kidney Diseases, and the Duke Cancer Institute. Th CPPOP study acknowledge support from the UCSF California Preterm Birth Initiative. The iLiNS-DYAD-M trial acknowledge the support by a grant to the University of California, Davis from The Bill & Melinda Gates Foundation [OPP49817] and a grant to the University of California, Davis from the Office of Health, Infectious Diseases, and Nutrition, Bureau for Global Health, U.S. Agency for International Development (USAID) through the Food and Nutrition Technical Assistance III Project (FANTA). GARBH -Ini cohort acknowledges the support by IUAC for extending Q-ICPMS under National Geochronology Facility funded by Ministry of Earth Sciences (MoES/P.O. (Seismic)8(09)-Geochron/2012). MDIG, AMANHI, GAPPS and INTERBIO cohorts acknowledge the support by The Bill & Melinda Gates Foundation.

