## Supplementary material for "Association of maternal prenatal copper concentration with gestational duration and preterm birth: a multi-country meta-analysis": Copper Supplementary Text 82322.docx

#### Supplementary Text 1. Regression analysis weighted by inverse of sampling probabilities.

**Bangladesh (AMANHI)** is a biobank in a population-based cohort of 3,000 pregnant women enrolled before 19 weeks of gestation and followed up-to 42 days post-partum. The overarching goal is to facilitate discoveries of biomarkers of adverse pregnancy outcomes (maternal, fetal and neonatal health outcomes) as new and more feasible methods become available. An additional goal is to identify biological mechanisms underlying the causes of the adverse outcomes including preeclampsia, spontaneous preterm birth (sPTB), stillbirth and intrauterine growth restrictions (IUGR) to create a platform to generate new approaches to treatment and prevention(26). All pregnant women were identified through pregnancy surveillance conducted by making home visits every 2 months by trained community health workers (CHWs). Pregnancies were confirmed via strip–based pregnancy tests and dated through ultrasound scans carried out by trained ultrasonologists before 19 weeks of gestation. The biobank contains maternal blood and urine specimens collected two times during pregnancy (8-19 weeks and 24-28 weeks or 32-36 weeks of gestation) and once during postpartum period (Day 42 postpartum) as well as delivery samples. Trained phlebotomists obtained maternal and umbilical cord blood samples and generated aliquots of serum, plasma, and buffy coats for storage. We also have collected and stored maternal urine, placental samples, umbilical cord blood, tissue, and membrane. When cord blood collection was not possible, saliva was collected from newborns. In addition, we collected infant blood at 12 months of age and paternal saliva samples. All samples were processed and stored in -80°C freezers. CHWs collected detailed phenotypic and epidemiological data from the pregnant women four times during pregnancy (at 8-19 weeks, 24-28 weeks, 32-36 weeks, and 38-40 weeks of gestation), at delivery and twice during postpartum period (<7 days and at 42 days). The study started in July 2014 and ended in April 2018.

**Bangladesh (GAPPS)** is a population based prospective cohort study of pregnant women with an objective to establish infrastructure for researchers to conduct preterm birth and pregnancy related research. The study enrolled 4220 pregnant women from Matlab district over three years (2015-2017). The research site enrolled women early in pregnancy and collected information and biological specimens during their pregnancy and delivery. Gestational age was estimated by standardized ultrasound method. All biological samples were stored immediately in the local biorepository using the standardized protocol developed by GAPPS domestic biorepository. The study was approved by the research ethics committee of the Matlab Health Research Center at the International Center for Diarrheal Disease Research, Bangladesh.

**Bangladesh (MDIG) study** is a randomized intervention trial of vitamin D supplementation during pregnancy and lactation (23). Participants were generally healthy pregnant women between 17 and 24 weeks of gestation that were enrolled between March 2014 and September 2015 at the Maternal and Child Health Training Institute in Dhaka, Bangladesh. Gestational age was estimated by ultrasound or LMP or both. The study was approved by research ethics committees at the Hospital for Sick Children at Toronto and the International Center for Diarrheal Disease Research, Bangladesh (icddr,b).

**Brazil, Kenya, Pakistan, South Africa, Thailand and UK (Interbio)** is a multicenter, population-based research initiative coordinated by the University of Oxford to assess human growth, neurodevelopment and associated behaviors from early pregnancy to 2 years of age (28). The Interbio study was conducted between February 2012 and June 2018 at sites: Pelotas (Brazil), Nairobi (Kenya), Karachi (Pakistan), Soweto (South Africa), Mae Sot (Thailand) and Oxford (UK). Gestational age was determined by standardized ultrasound method. The studies were approved by regional ethics or institutional research boards and by the Institutional review board at the University of Oxford.

**Malawi (iLiNS-DYAD)** is a randomized, controlled, partially blinded, parallel-group intervention trial known as the International Lipid-based Nutrient Supplements DYAD trial which was designed to study the health impacts of lipid-based nutrient supplements during pregnancy and lactation (22). The study was conducted in 2 hospitals and 2 health centers in a rural area in Mangochi district between 2011-2012. Participants of the Malawi cohort were enrolled from four health facilities that covered mostly one continuous area near Lake Malawi. Lungwena, Malindi, and Mangochi subsites are along the banks of Lake Malawi and close to Namizimu forest reserve. Gestational age was determined using the ultrasound method. The ethical clearance for the study was granted by the University of Malawi College of Medicine Research and Ethics Committee (COMREC) and the ethics committee at Tampere University Hospital District, Finland.

**Pakistan (AMANHI)** is a population-based biorepository with the aim of collecting cause-specific biosamples on maternal and neonatal mortality, and stillbirths from a well-characterized cohort of pregnant women(26). The biobank enrolled 2,500 pregnant women from 2014 to 2018 with the last pregnancy outcome occurring in January 2019, after an ultrasound in early pregnancy (<20 weeks) to confirm the gestational age. Women were also visited thrice at 24-28, 32-36 and 38-42 weeks gestation during pregnancy, and 0-6 days and 42-59 days after birth to measure blood pressure, test urine for proteinuria, and ascertain reported morbidity since the previous visit. The outcome of each identified pregnancy, whether abortion, stillbirth or live birth, was carefully documented and verbal autopsies were conducted with appropriate respondents in case of maternal deaths, stillbirths or neonatal deaths. Subsequently, all live born babies were assessed for neuromuscular, physical and feeding maturity as well as neonatal anthropometry (including baby’s weight and foot length) within 72 hours of birth up until 4-5 years of age, using harmonized procedures. Blood and urine samples were collected from each woman at three time points: enrolment (<20 weeks), at either 24-28 weeks or 32-36 weeks gestation, and at 42 days postpartum. These samples were processed and stored at -80ºC in multiple aliquots. At delivery of a still- or live birth, maternal stool, umbilical cord blood and placental were collected, processed and stored within 30 minutes of birth. At the 42 days postpartum visit, maternal blood and urine, infant stool and paternal saliva samples were also collected for processing and storage. Infant saliva was collected if cord blood could not be obtained. Study protocols for enrolment, visits, ultrasound scans, sample collection, processing and storage were implemented by highly trained and motivated staff. Community members were found to be very cooperative and supportive of the study. The study was approved by the regional ethics review committee.

**Tanzania (AMANHI)** Tanzania (Pemba) is one of the AMANHI sites with bio-banked biological samples from a cohort of 4501 pregnant women and their children. The overall objective of all participating sites in the AMANHI study was similar, and all the SOP implemented for sample collection and processing were harmonized across sites(26). The study was conducted in Pemba Island of the Zanzibar archipelago with an overall population of around 432000 and approximately 82,000 households. Two districts of the island were selected for the study where pregnancy surveillance was conducted every 2 months to identify pregnant women. Consent was obtained for confirmation of pregnancy by urine strip tests, thereafter gestational age was confirmed with routine ultrasound methods. All women between 8-19 weeks of gestation were consented and then enrolled in the study. Blood and urine samples were collected from the enrolled mothers at the time of enrollment and in either during 24 – 28 weeks or 32 – 36 weeks of gestation and 42 days postpartum. At delivery in addition to cord blood, tissue samples from placenta, membrane cord were collected within 30-60 minutes of delivery. All samples were processed as per harmonized SOP and stored in the biobank at -80^0^ C. Saliva samples from the father and fecal samples from the mother and infant were also collected after delivery. For collecting the epidemiological data study team visited all the mothers during pregnancy and after delivery. Additional information on delivery was obtained from the hospital delivery records filled in by the physician in-charge. A in house designed AMANHI biobank LMIS software was used by all AMANHI sites for recording the collection, processing and storage information of the biospecimens. Stringent quality control checks were implemented for data consistency and sample quality during the study. The study started in June 2014 and the last postnatal follow-up sample from the mother was collected in December 2018.

**UK (Liverpool) was started** in April 2012 and ended in December 2017. The study entitled “The development of novel biomarkers for prediction of preterm labor in a high-risk population”. This study enrolled a total of 541 pregnant women with singleton pregnancies at the Liverpool Women’s Hospital, UK. Two populations of women were targeted for recruitment. The first was a ‘low risk’ group who were parous women with all previous births at term. The second was a ‘high risk’ group of women who had a history of spontaneous preterm birth or preterm prelabour rupture of membranes under 34 weeks gestation. Gestational age was determined using the ultrasound method. Samples were obtained between 15^+0^ and 23^+0^ weeks gestation. Gestational age was determined using the ultrasound method. Samples were obtained between 15^th^ and 23^rd^ weeks of gestation. The study was approved by the Institutional Review board at the University of Liverpool.

**USA, CA(CPPOP)** is a nested case-control sampling study drawn from a population-based cohort of 757,853 singleton live births in the state of California (24). Women with nonfasted serum samples banked by the California biobank program from July 2009 through December 2010 were enrolled in the study. Gestational age was determined by ultrasound or LMP or both. Methods and protocols for the study were approved by the Committee for the Protection of Human Subjects within the Health and Human Services Agency of the State of California, the Institutional Review Board of Stanford University, and the Institutional Review Board of the University of California San Francisco.

**USA, NC (NEST)** is a perinatal cohort study of more than 2000 pregnant and their offspring mounted to investigate the role of environmental exposures and nutrition in utero on the shifts in the epigenome of newborns (25), from which we nested a case control study. The study participants were a birth cohort from women who received prenatal care in the Duke/Durham region health care system in Durham, NC between 2005 and 2009. Ultrasound is used to determine the gestational age. The study was approved by the Institutional Review Board of Duke University, North Carolina.

**Vietnam (PBB)** is an observational study in Ho Chi Minh city to evaluate biomarkers for spontaneous preterm birth in partnership with Sera Prognostics, OUCRU and the Gates Foundation. This is a prospective hospital-based convenience sampling of 4800 women between September 2016 through August 2018 who have antenatal care and plan to deliver at Tu Du hospital and were between 19^+0^ to 22^+6^ days of gestation when samples were taken. Gestational age was determined using ultrasound. The study was approved by the Institutional review board at the Tu Du Hospital and the University of Oxford (OXTREC 28-16).

**Zambia (GAPPS)** is a population based prospective cohort study of pregnant women with an objective to establish infrastructure for researchers to conduct preterm birth and pregnancy related research. The study enrolled 2000 pregnant women from Lusaka district over three years (2015-2017). The research site enrolled women early in pregnancy and collect information and biological specimens during their pregnancy and delivery. Gestational age was estimated by ultrasound at gestational weeks 16-22. All biological samples were stored immediately in the local biorepository using the standardized protocol developed by GAPPS domestic biorepository. The study was approved by the regional ethics review board.

**India (THSTI)** is a hospital-based pregnancy cohort **(GARBH – Ini**: Interdisciplinary Group for Advanced Research on Birth Outcomes - DBT India Initiative), initiated in May 2015 at the civil hospital in Gurugram district, Haryana, India. It is a part of collaborative interdisciplinary program coordinated by Translational Health Science and Technology Institute (THSTI), Faridabad, India, supported by the Department of Biotechnology (DBT) Government of India. The program is a unique collaboration between research institutes [THSTI, Regional Centre for Biotechnology (RCB), National Institute of Biomedical Genomics (NIBMG)] and Gurugram Civil Hospital, Haryana. Gurugram Civil Hospital is the primary clinical site with Safdarjung Hospital, New Delhi as the referral center. The study was approved by institutional ethics committees, both at the hospital site and at THSTI. It is an ongoing cohort study comprising >8000 pregnant women from rural and semi-urban population, enrolled within 20 weeks of gestation and followed until delivery and postpartum (Bhatnagar et al 2019). In brief, pregnant women are approached in the antenatal clinic, by the study nurses, and screened based on their last menstrual period. The women are enrolled in the study after a dating ultrasound confirming a POG of <20 weeks. With written informed consent, socio-demographic information, longitudinal ultrasonographic images and bio-specimen are collected and stored at respective repositories, to decipher the epidemiological, clinical, and biological correlates of adverse maternal and child health outcomes primarily Preterm Birth (PTB). The primary hypothesis is “time-series data on a large set of variables - including clinical, environmental, nutritional, genomic, epigenomic, metagenomic, and proteomic - collected across pregnancy will help in stratifying women into defined risk groups for PTB”. To comprehend population specific, multiple micro/macro nutrient deficiencies associated with PTB, these participants are being profiled for various nutritional parameters, and serum copper levels in early pregnancy is one of them. The present article comprises information about serum copper levels on a subset of participants (N=507, bio specimen collected from February 2016 to January 2017. The eligibility criteria for the subset of participants were singleton pregnant women enrolled before 20 weeks of gestation, with availability of maternal sera at the same time points, and delivery outcomes documented at the hospital site.

#### Supplementary Text 2. Protocol for Copper analysis in plasma and serum samples

**Sample Preparation**

All serum and plasma samples were randomized, before analysis these were thawed on ice for 30 minutes followed by sonication in an ultrasonic bath (Fisher Scientific, CPX1800) for 5 minutes in order to mix the samples. During these processes, the sample vials remained sealed closed in order to prevent dilution of the sample caused by the premature opening of the sample vials before they reach room temperature. An acid digestion was performed on the samples, 50 µL of plasma or serum sample, as well as the quality control serum was transferred to 15 mL metal free vials (VWR) using clear pipette tips and a calibrated electronic micropipette (Eppendorf). Both the samples and the quality control serum were mixed briefly immediately prior to transferring the aliquot using a mini vortex mixer (VWR) to promote better sampling of a nonhomogeneous sample matrix. The quality control serum used was obtained from UTAK Laboratories, Inc. and included normal range trace elements which was reconstituted according to manufacturer instructions.

50 µL of the internal standard mixture containing 500 ppb of Sc, In, Y and Te in 0.5 M nitric acid (High Purity Standards) was then added with a repetition pipette (Eppendorf) followed by 200 µL of concentrated trace metal grade nitric acid (Sigma-Aldrich; Fisher Scientific). The samples were placed in a dry bath with no more than 3 cm of the tubes immersed in the heating block holes in order to allow reflux of the sample. For this, aluminum foil was inserted into the block holes.

The samples were heated at 85 oC for one hour, vented, heated at 95 oC for 1.5 hours. Then 100 µl of trace metal grade hydrogen peroxide (Sigma-Aldrich) was added with a repetition pipette after the samples cooled for 5 minutes outside the heating block. Then the samples were returned to the heating block another 30 minutes at 95 oC. Once the digestion was completed, the final volume was brought up to 2.5 ml with doubly deionized water and mixed using a vortex mixer.

**Calibration Preparation**

The external calibration method with internal standard in-samples was used. For this the following points were used: 0, 0.5, 1, 2, 5, 10 and 25 ppb. The standards were prepared in 0.5 M trace metal grade nitric acid from a stock solution containing a mixture of elements of interest at 10 ppm (SPEX CertiPrepTM).

**ICP-MS analysis**

An Agilent 7700 ICP-MS sytem was used for the copper quantification. The collision/reaction cell was used with helium to eliminate polyatomic interferences. A micro concentric nebulizer was used, with a double pass Scott spray chamber at 2 oC, connected to a standard 2 mm torch and with platinum sampler and skimmer cones. The system was operated with the Agilent Mass Hunter ICP-MS software. Copper was quantified at the 63Cu istotope with 0.1 s of integration time and with 89Y as internal standard. A typical batch of 120 samples plus quality control samples consisted in 2 instrumental replicates of the digested UTAK CRM, one at the beginning and one at the end of the run, three standards of 1 and 2 ppb at the middle and end of the sequence, and 2 biological replicates of the in-house QC serum pool in random order, and three technical replicates of digestion blanks every 45-50 samples.

### Supplementary Tables

#### STable 1. Study characteristics of participant cohorts

| **Site** | **Location** | **Study design, sample collection** | **Year** | **Data sharing format with CCHMC** | **GA estimation method** | **Type of Sample** |
| --- | --- | --- | --- | --- | --- | --- |
| Bangladesh (AMANHI) | Sylhet | Population based, random | 2012-2016 | Case:Control (1:1) | Ultrasound | Plasma |
| Bangladesh (GAPPS) | Matlab | Population based, random | 2015-2017 | Case:Control (1:2) | Ultrasound | Serum |
| Bangladesh (MDIG) | Dhaka | Hospital based, intervention trial | 2014-2015 | Case:Control (1:2) | Ultrasound & LMP | Serum |
| Brazil (INTERBIO) | Pelotas | Hospital based, random | 2009-2014 | Random | Ultrasound | Plasma |
| India (THSTI) | Gurugram | Hospital based, random | 2016-2017 | Random | Ultrasound | Plasma |
| Kenya (INTERBIO) | Nairobi | Hospital based, random | 2009-2014 | Random | Ultrasound | Plasma |
| Malawi (iLiNS-DYAD) | Mangochi | Hospital based, intervention trial | 2011-2015 | Random | Ultrasound | Plasma |
| Pakistan (AMANHI) | Karachi | Population based, random | 2012-2016 | Case:Control (1:2) | Ultrasound | Serum |
| Pakistan (INTERBIO) | Karachi | Hospital based, random | 2009-2014 | Random | Ultrasound | Plasma |
| South Africa (INTERBIO) | Johannesburg | Hospital based, random | 2009-2014 | Random | Ultrasound | Plasma |
| Tanzania (AMANHI) | Pemba | Population based, random | 2009-2016 | Case:Control (1:2) | Ultrasound | Plasma |
| Thailand (INTERBIO) | Mae Sot | Hospital based, random | 2009-2014 | Random | Ultrasound | Plasma |
| UK (INTERBIO) | Oxford | Hospital based, random | 2009-2014 | Random | Ultrasound | Plasma |
| UK (LIVERPOOL) | Liverpool | Hospital based, targeted recruitment | 2016-2017 | Random | Ultrasound | Plasma |
| USA, CA (CPPOP) | San Francisco | Hospital based, | 2009-2010 | Case:Control (1:1) | Ultrasound & LMP | Serum |
| USA, NC (NEST) | Durham | Hospital based, random | 2005-2009 | Case:Control (1:2) | Ultrasound | Plasma |
| Vietnam (PBB) | Ho Chi Minh City | Hospital based, random | 2016-2018 | Case:Control (1:2) | Ultrasound & LMP | Serum |
| Zambia (GAPPS) | Lusaka | Hospital based, random | 2015-2017 | Random | Ultrasound | Serum |

#### STable 2. Phenotype data requested from sites

| **Variable** | **Required** | **Desired** |
| --- | --- | --- |
| ***Baseline characteristics*** |  |  |
| Gestational Age (at time of sample collection) | x |  |
| Maternal Age |  | x |
| Maternal race |  | x |
| Maternal ethnicity |  | x |
| ***Birth history*** |  |  |
| Gravidity (# of pregnancies) |  | x |
| Parity (# of births) |  | x |
| # prior PTB |  | x |
| # prior stillbirth |  | x |
| ***Pre-pregnancy BMI*** |  |  |
| Height/weight at visit | x |  |
| Pre-pregnancy weight (if available) | x |  |
| ***Exposures*** |  |  |
| Smoking during pregnancy |  | x |
| Alcohol during pregnancy |  | x |
| Substance use during pregnancy |  | x |
| ***Delivery outcomes*** |  |  |
| Delivery date |  | x |
| Gender | x |  |
| Gestational age at delivery | x |  |
| Birth weight | x |  |
| Spontaeous versus indicated delivery (if available) | x |  |
| Infant/fetus vital status (live birth) | x |  |
| ***Conditions*** |  |  |
| Chorioamnionitis |  | x |
| Hypertensive disorder |  | x |
| Preeclampsia |  | x |
| Gestational diabetes |  | x |
| Other specified conditions |  | x |

#### STable 3. Prevalence, case/control ratios and sampling probabilities

| **Case/control sites** | **prevalence** | **case/control ratio** | **case/control sampling probability** |
| --- | --- | --- | --- |
| Bangladesh (AMANHI) | 0.12 | 1 | 7.33 |
| Bangladesh (GAPPS) | 0.11 | 0.5 | 4.05 |
| Bangladesh (MDIG) | 0.09 | 0.52 | 5.24 |
| Pakistan (AMANHI) | 0.14 | 0.5 | 3.06 |
| Tanzania (AMANHI) | 0.05 | 0.5 | 9.54 |
| USA, California (CPPOP) | 0.09 | 1 | 10.09 |
| USA, North Carolina (NEST) | 0.11 | 0.49 | 3.95 |
| Vietnam (PBB) | 0.09 | 0.49 | 4.96 |

### Supplementary Figures

#### SFigure 1. Distribution of gestational duration of singleton live births from all sites

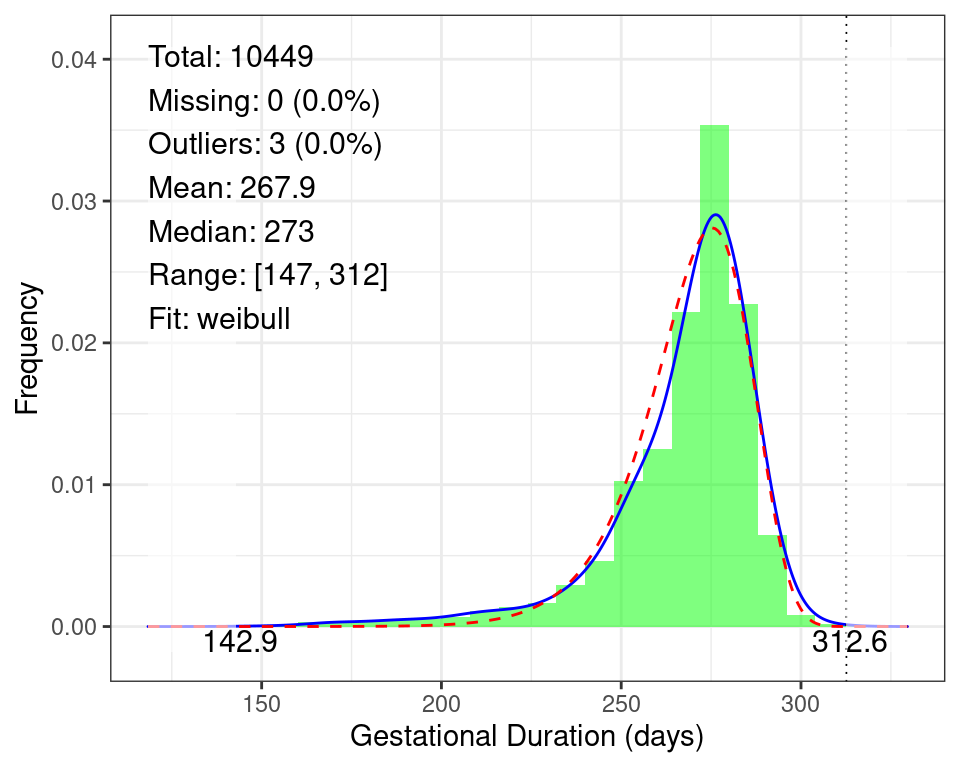

#### SFigure 2. Gestational duration in term and preterm deliveries by participating sites

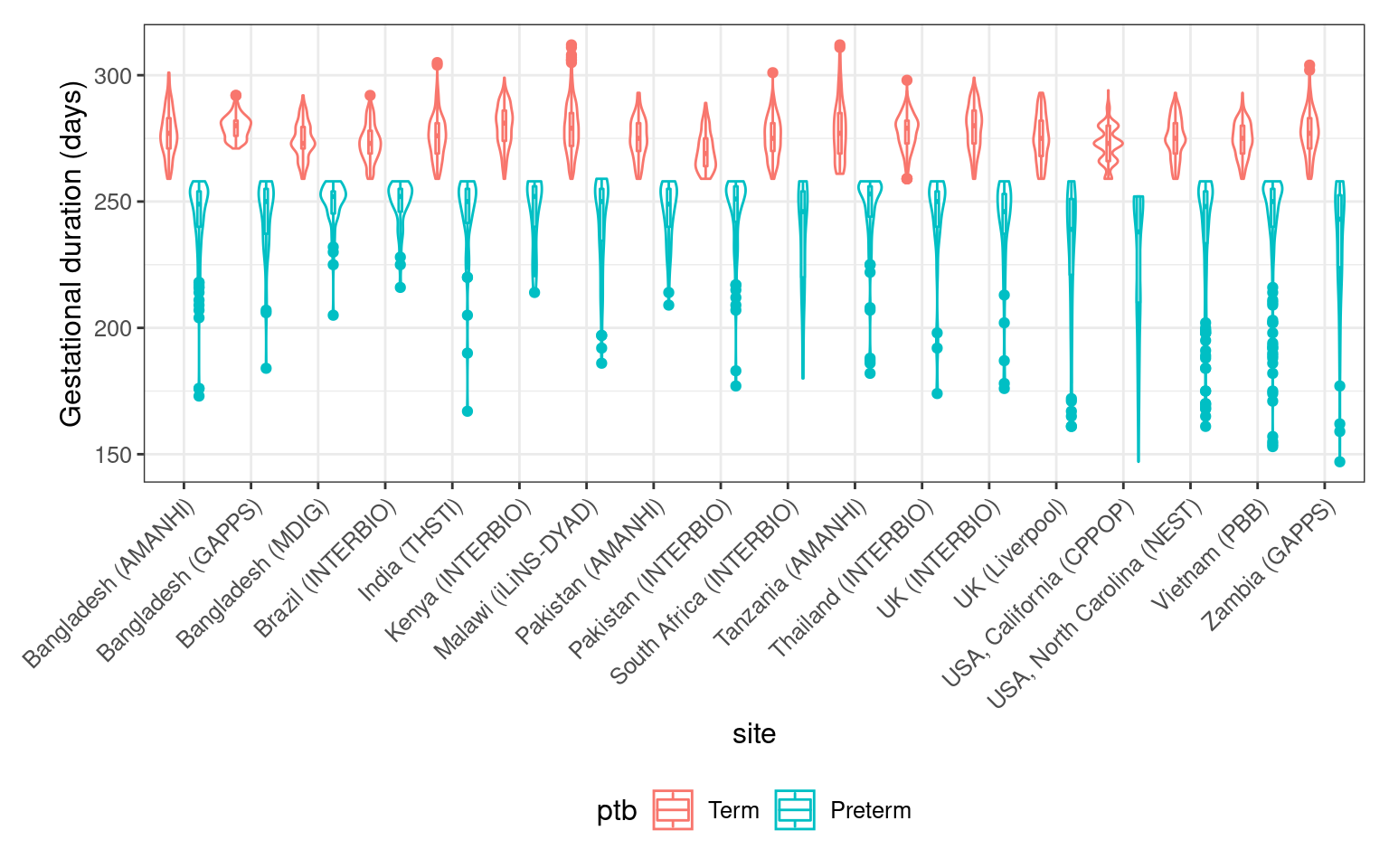

Violin plot illustrating the distributions of gestational days in term (gday ≥ 259 days) and preterm (gday < 259 days) deliveries from each site.

#### SFigure 3. Correlation of gestational duration and preterm birth with other covariates

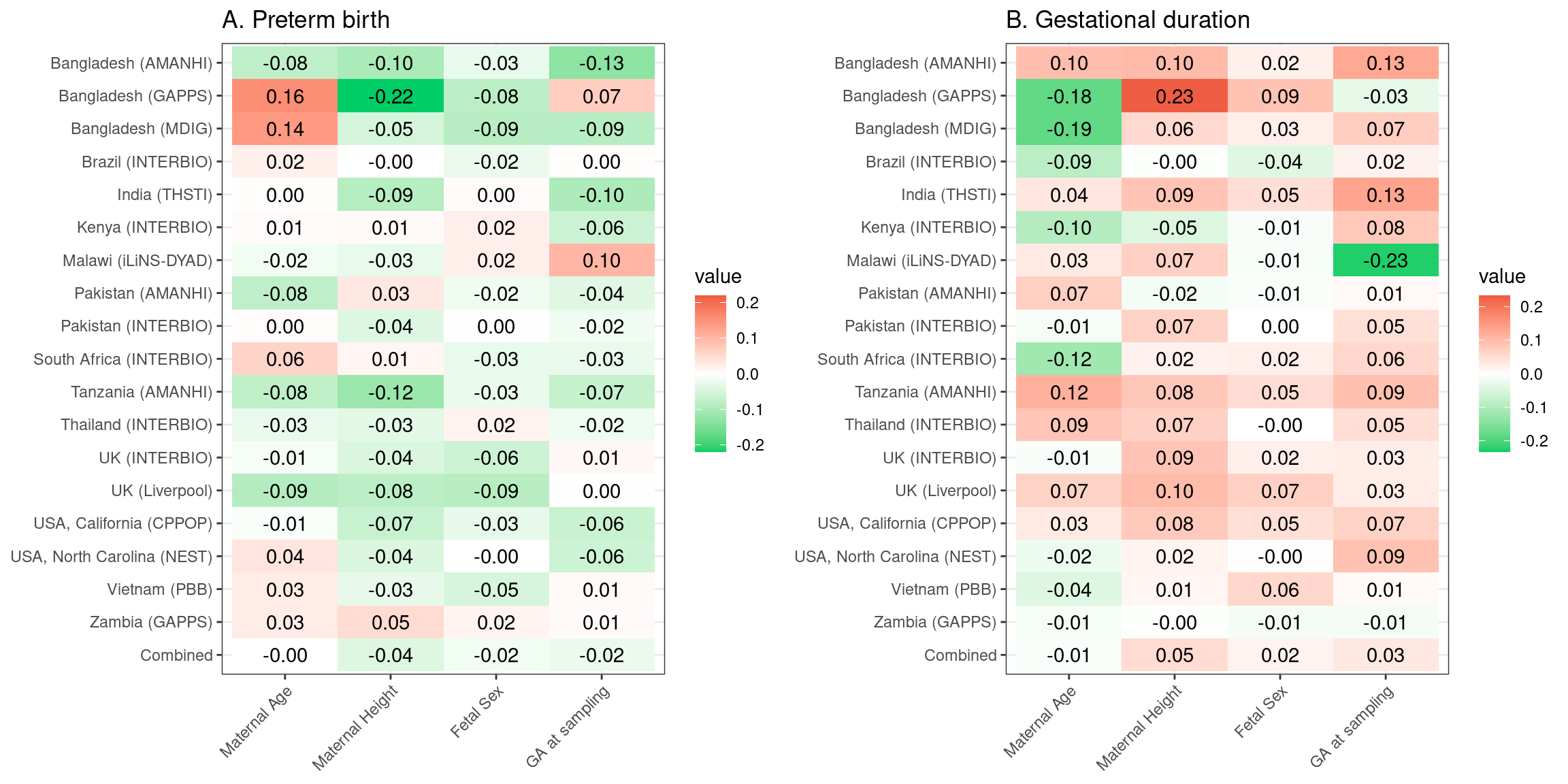

Heat maps illustrating the correlation of preterm birth (A) and gestational duration (B) with pregnancy covariates. Red shading indicates a positive correlation and green a negative correlation, with intensity reflecting the magnitude of the correlation.

#### SFigure 4. Distribution of maternal Cu concentration of samples from all sites

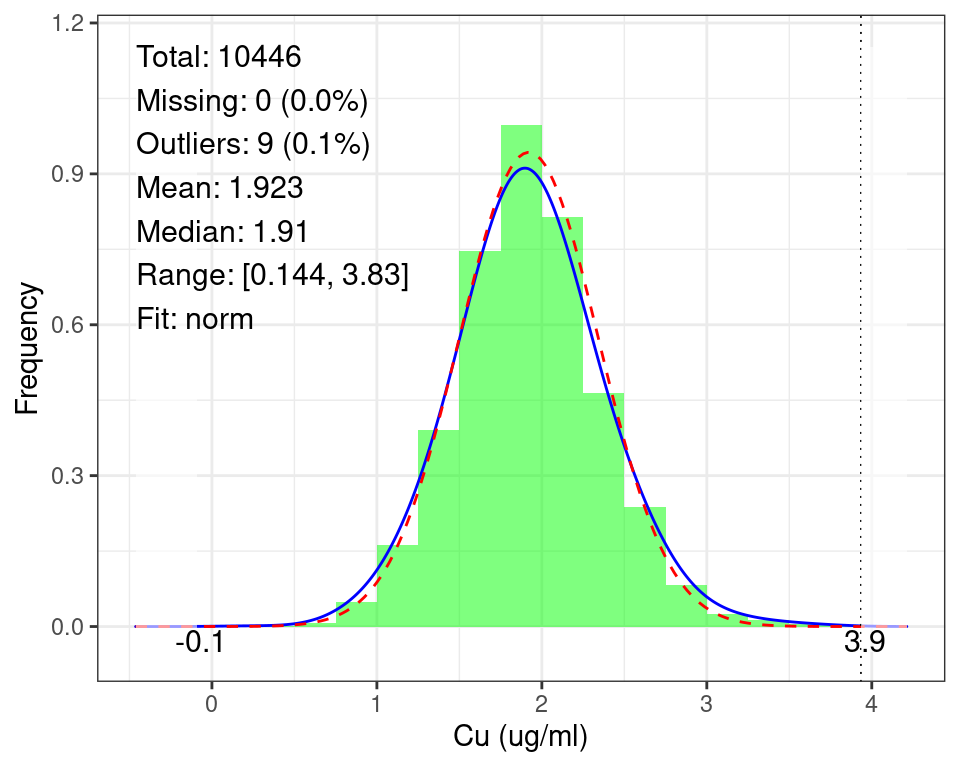

#### SFigure 5. Maternal Cu concentration by participating sites

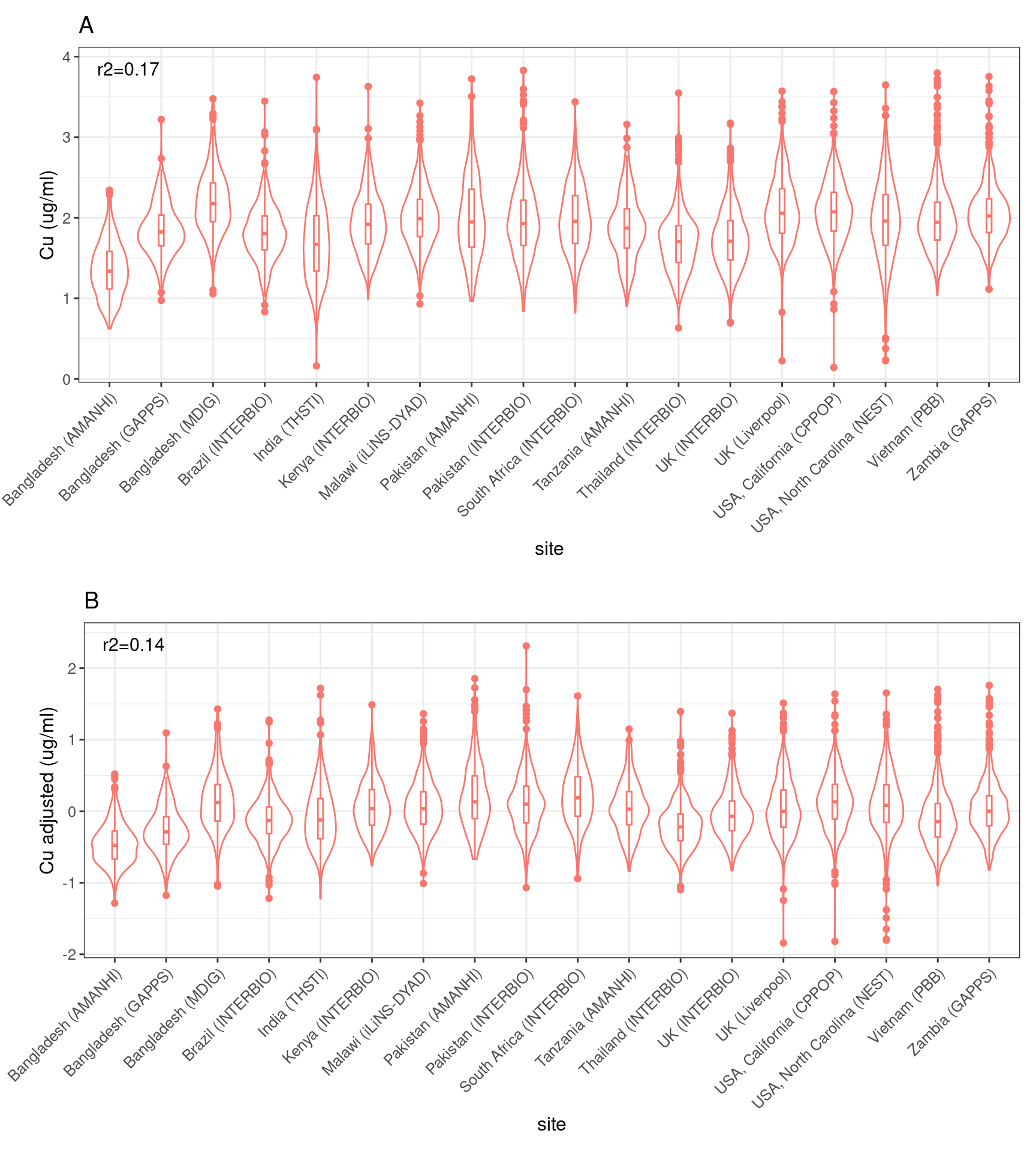

A. Raw Cu concentration: B. Cu concentration adjusted by gestational age at sampling

r2 is the variance in Cu concentration explained by sites.

#### SFigure 6. Maternal Cu concentration in term and preterm deliveries by participating sites

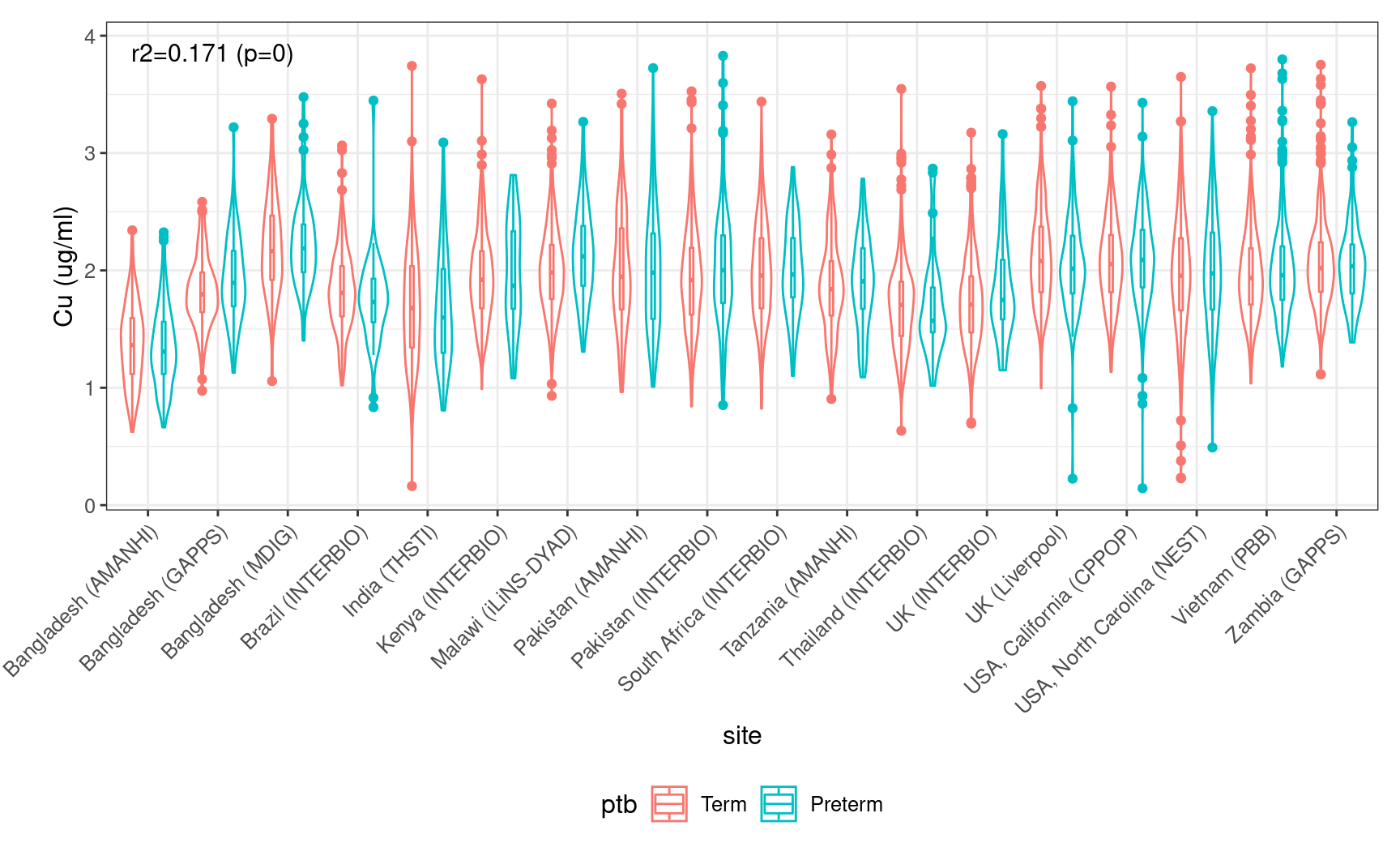

r2 is the variance in Cu concentration explained by sites.

#### SFigure 7. Maternal Cu concentration measured at different batches (colored by site)

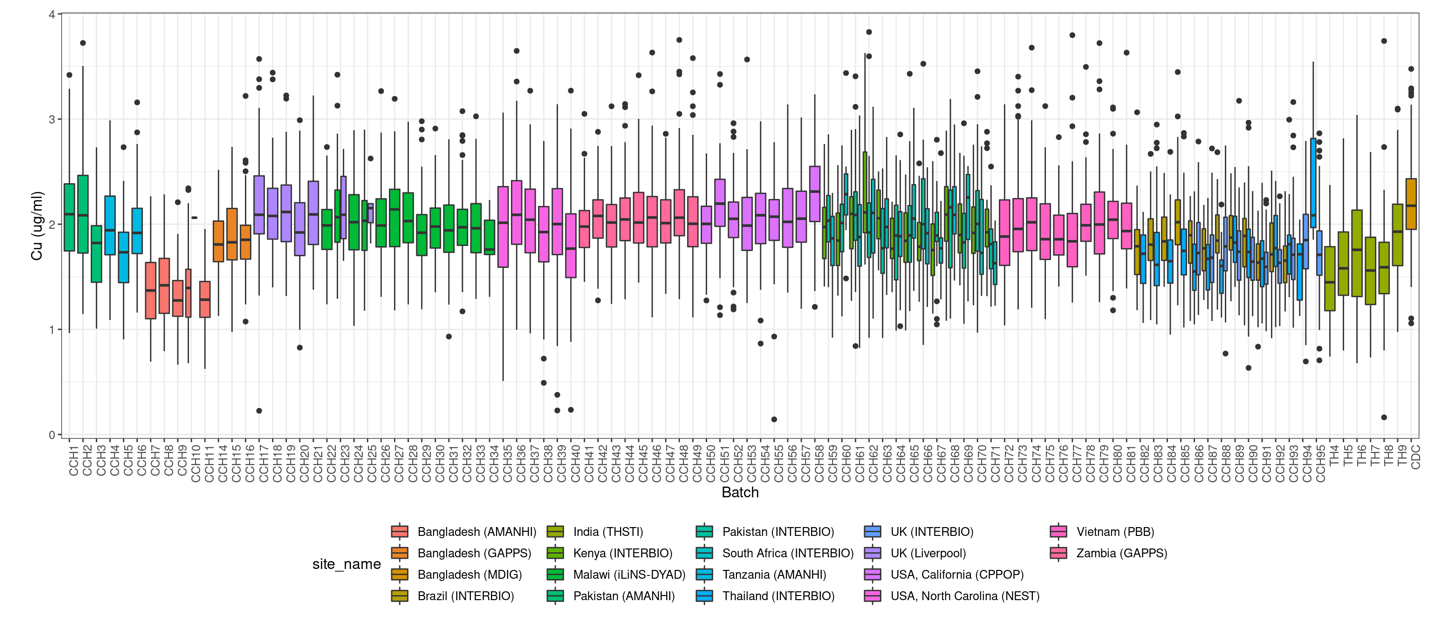

#### SFigure 8. Correlation of maternal Cu concentration with other covariates

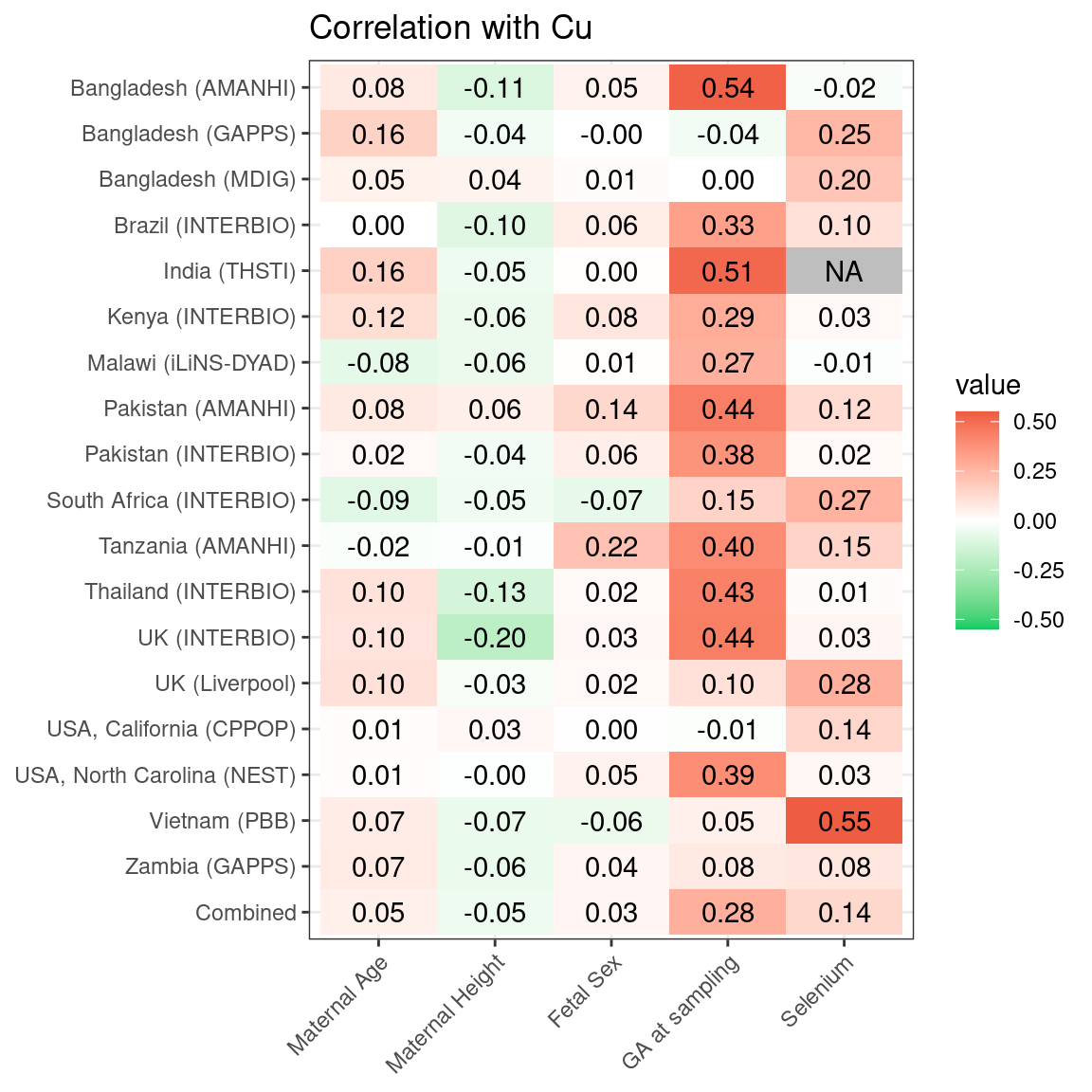

#### SFigure 9. Gestational age (weeks) at sample collection by sites

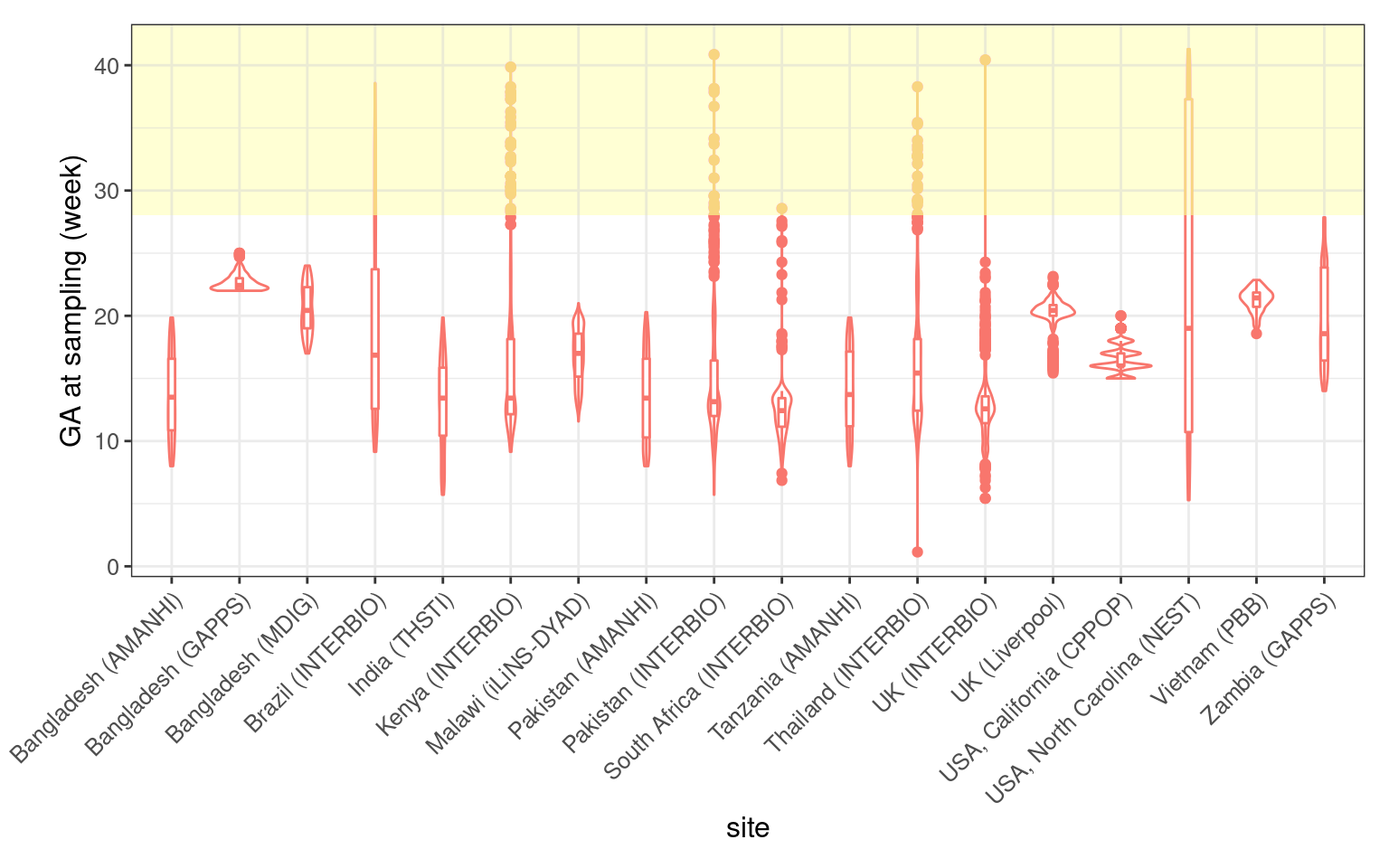

Violin plot illustrating the distribution of gestational age (weeks) at sample collection by sites. Yellow shaded region represents the samples collect after 2nd trimester (≥ 28 wks), which were excluded from the final association analysis.

#### SFigure 10. Association between adjusted Cu and gestational duration

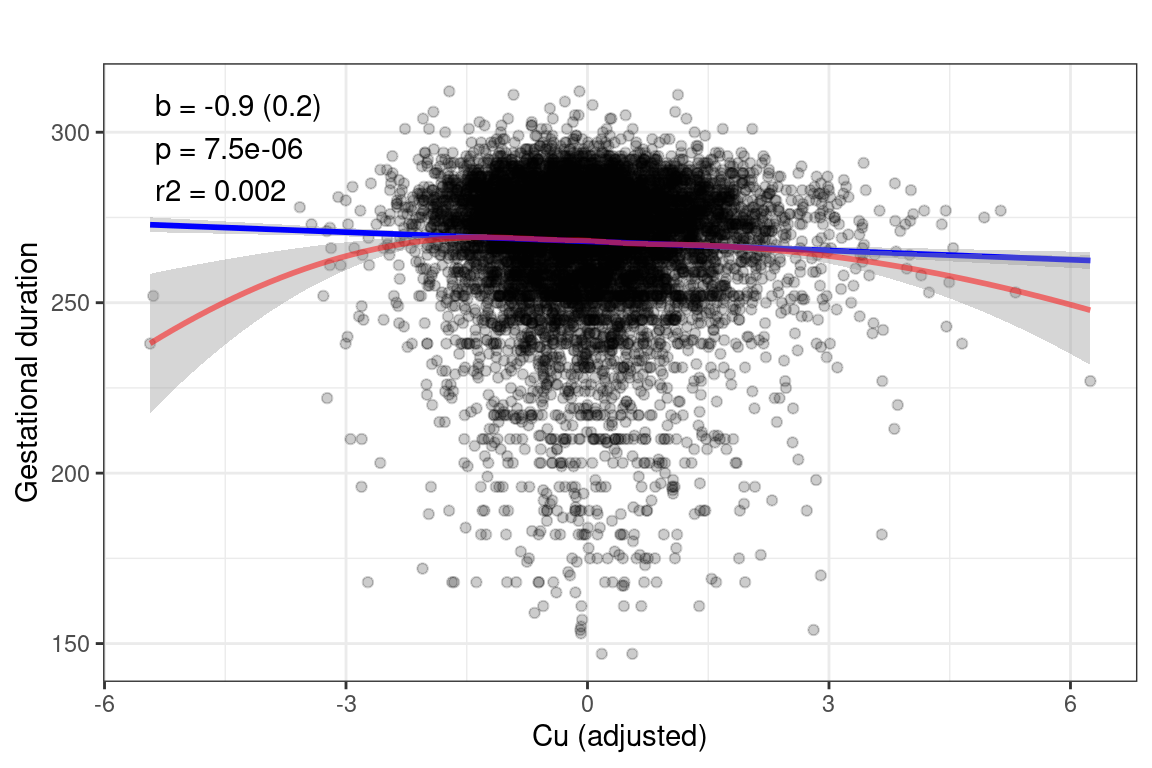

The linear regression (blue line) and LOESS fitting (red curve) between adjusted Cu (by gestational age at sample collection) and gestational duration. The adjusted Cu is scaled to sd=1.

b is the effect size estimate based on linear regression analysis of adjusted Cu against gestational duration, p is the associated p-value by t-test and r2 is the variance in gestational duration explained by adjusted Cu.

#### SFigure 11. Fraction of term, preterm births in quartiles of Cu concentration

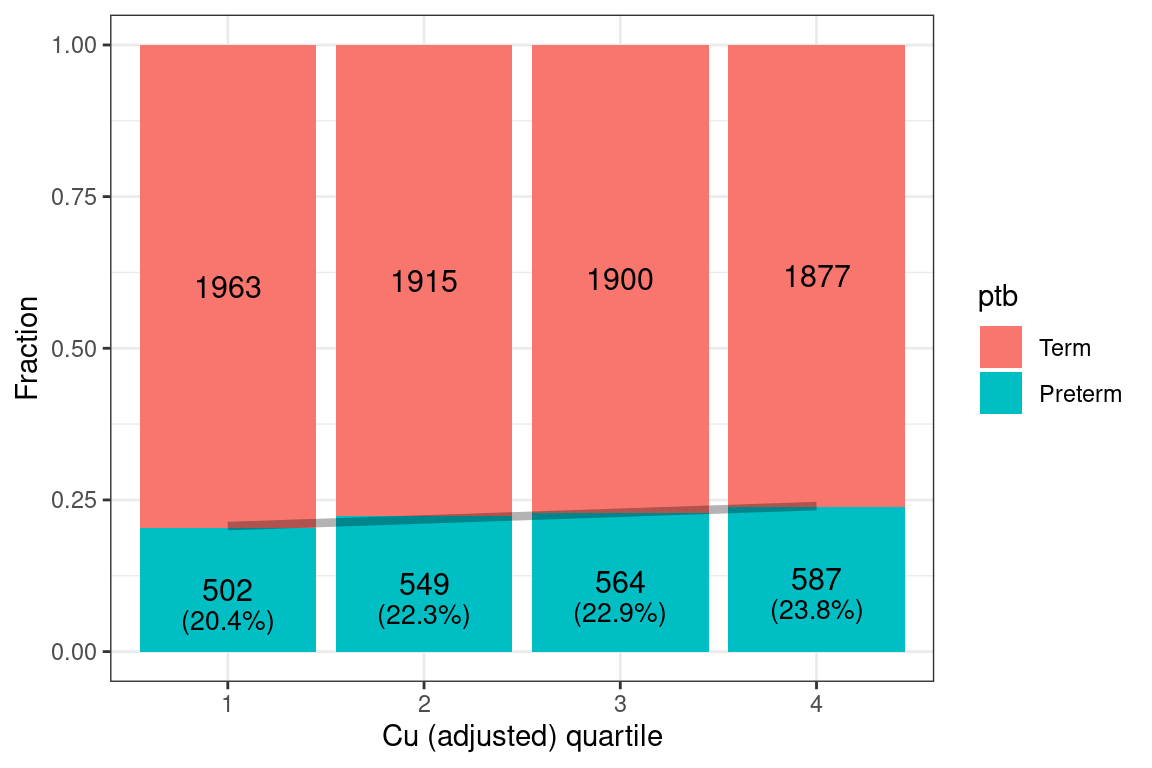

The shaded line shows the linear regression between quartile and preterm rates.

#### SFigure 12. Violin plots of key variables in Malawi subsites

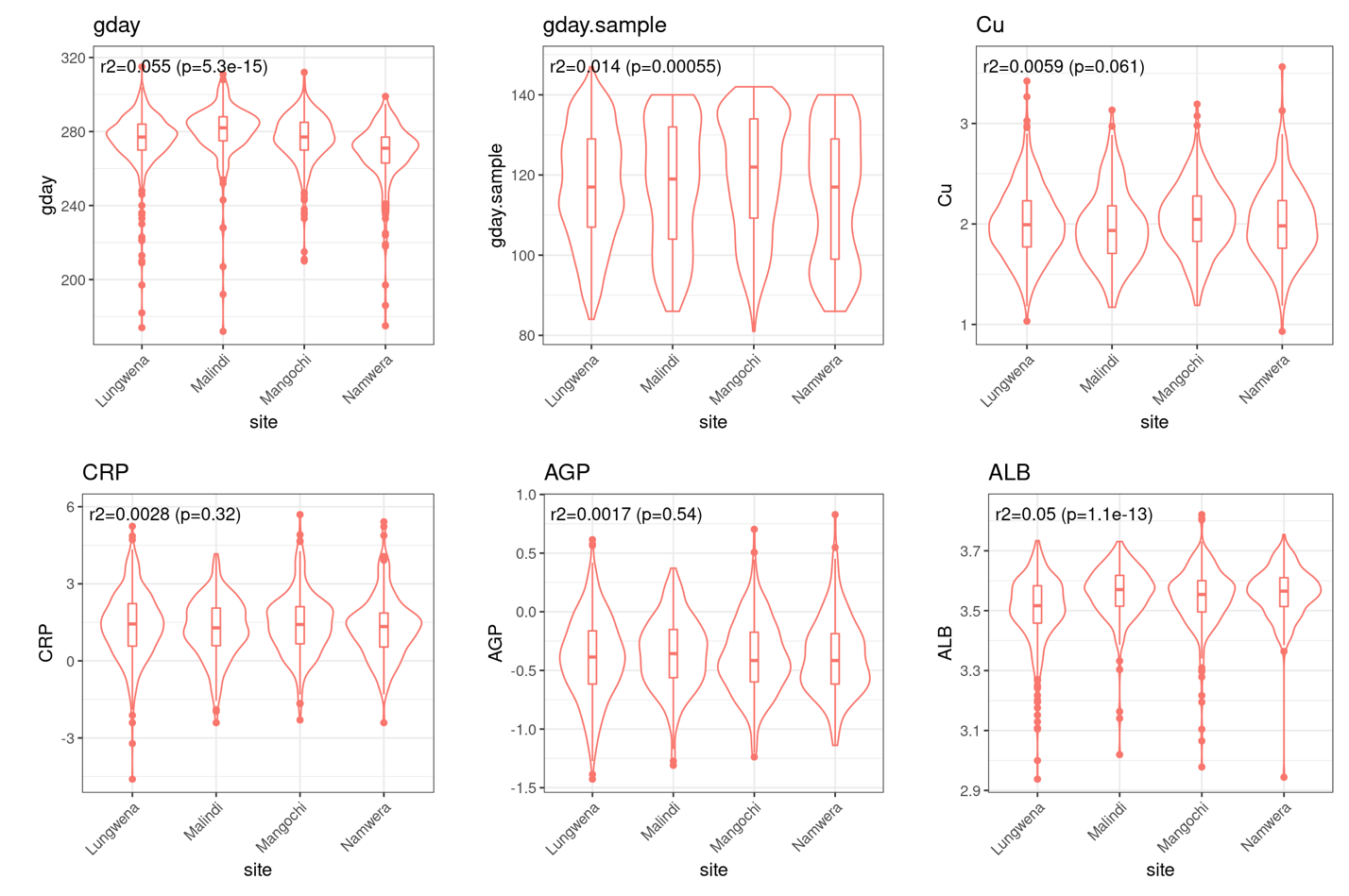

r2 is the variance of the dependent variable (y-axis) explained by sites (x-axis) and p is the associated p-value by ANOVA test.

#### SFigure 13. Association between gestational age at sampling and concentrations of maternal Cu and common analytes

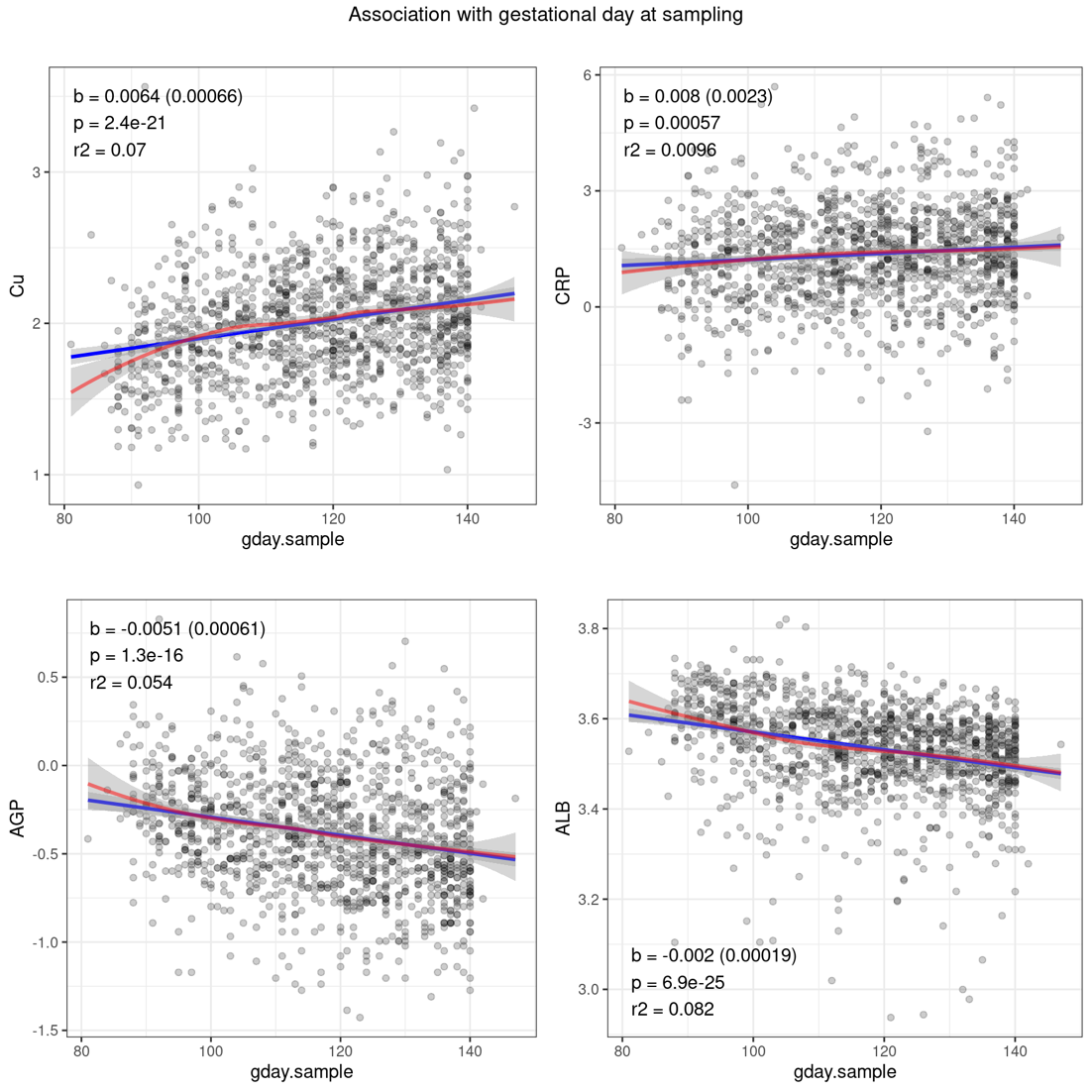

b is the effect size estimate based on linear regression analysis of gestational days at sampling (x-axis) against a bioanalyte (y-axis), p is the associated p-value by t-test and r2 is the variance in the bioanalyte explained by gestational days at sampling.

#### SFigure 14. Associations between Cu and analytes without or with adjustment of gestational age at sampling

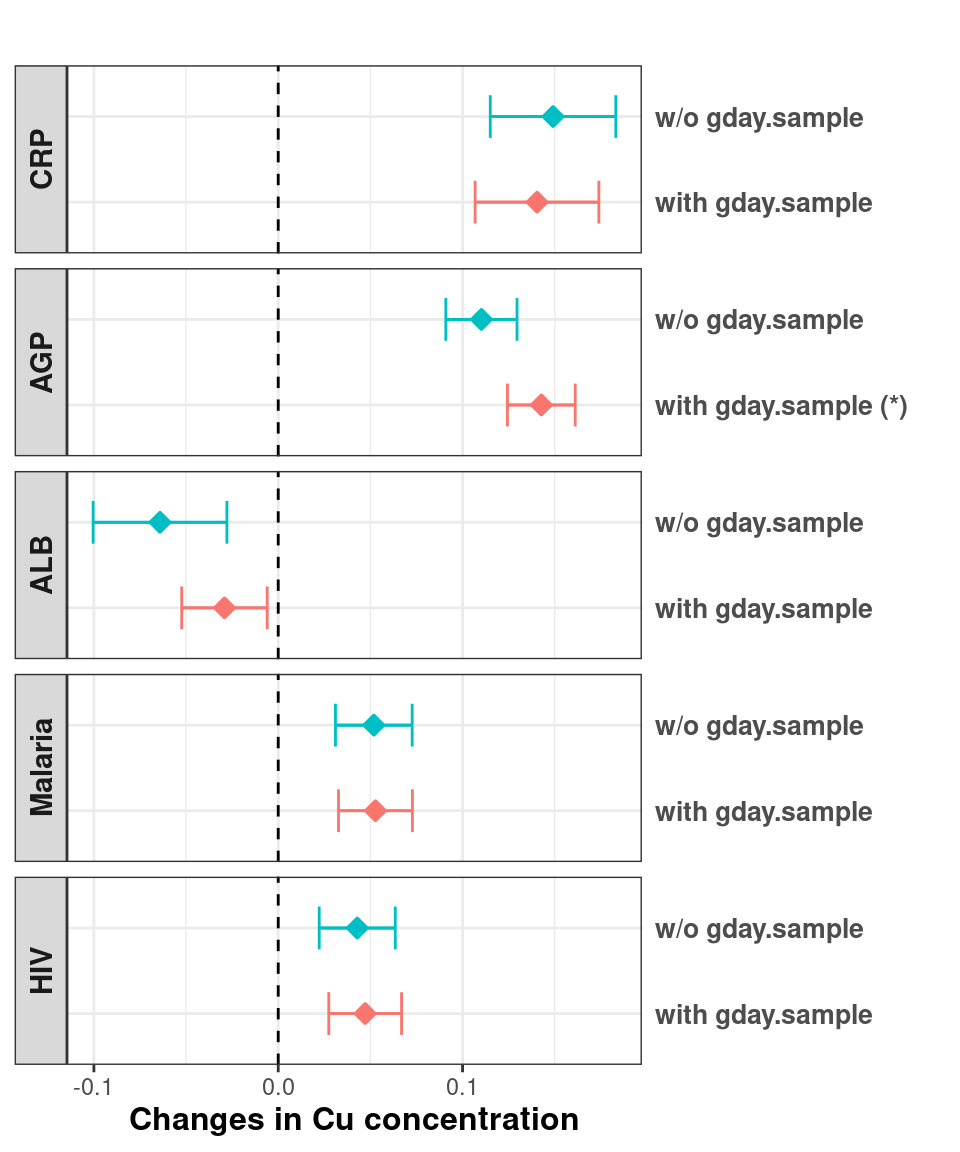

The estimated effects of different analytes (in sd), malaria, and HIV infections (log odds) on Cu concentration without (w/o) or with including gestational days at sampling (gday.sample) as a covariate.

#### SFigure 15. Estimated effect of Cu on PTB and gestational duration under different association models

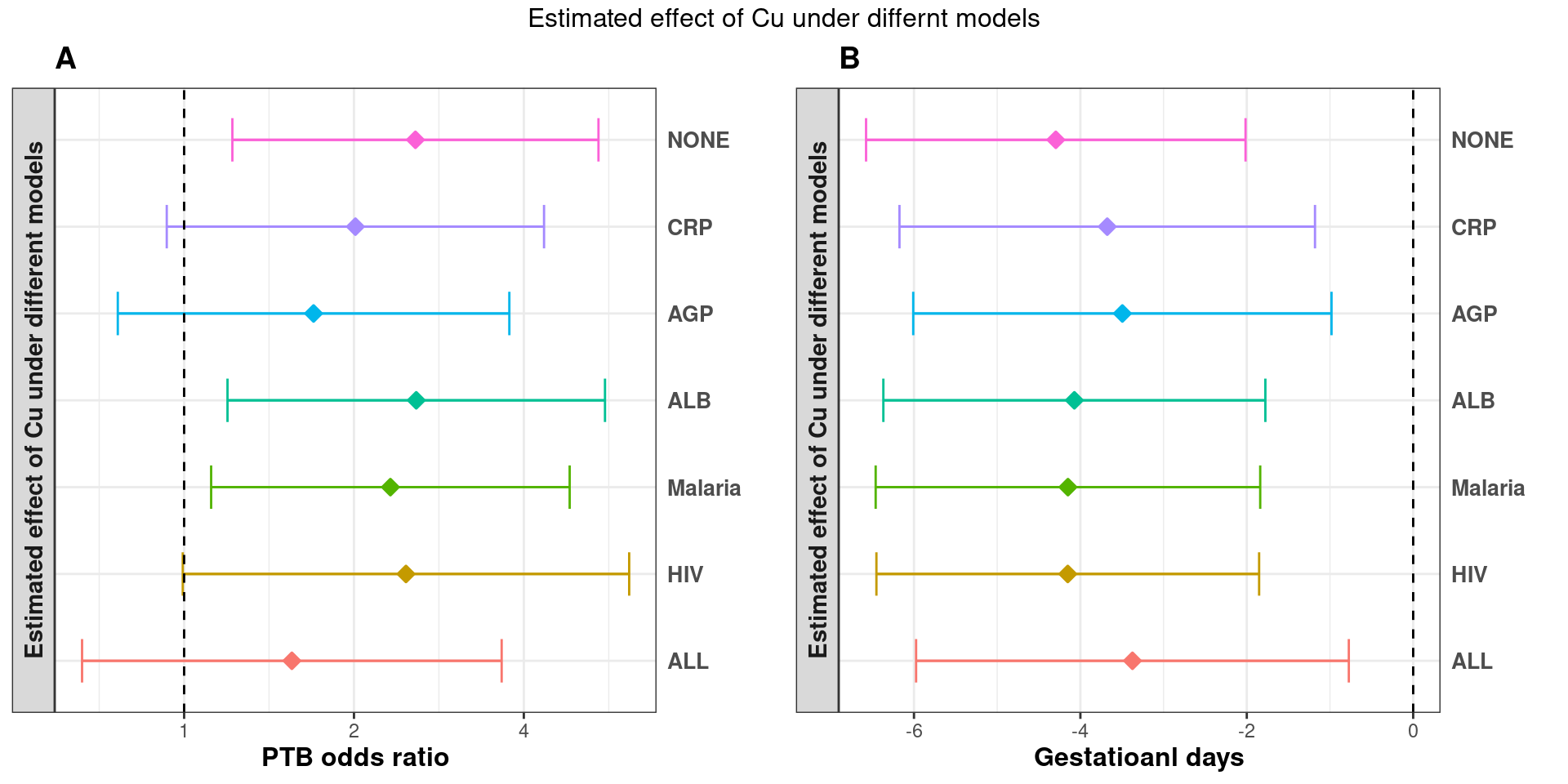

Estimated effect of Cu (per 1μg/ml) on PTB (A) and gestational days (B) using different regression models. NONE: none of the other APRs or infections was included as covariate; CRP, AGP, ALB, Malaria and HIV: individual APRs or infections was included as a covariate; ALL: all the APRs and infections were included together as covariates.
